# Antimicrobial resistance research in Singapore – mapping current trends and future perspectives

**DOI:** 10.1101/2023.11.28.23299149

**Authors:** Selina Poon, Dai Mei Goh, Astrid Khoo, Yueh Nuo Lin, Yee Sin Leo, Tau Hong Lee

## Abstract

Antimicrobial resistance (AMR) research is increasing globally, but its extent in Singapore is unclear. The aim of this study was to review the current research trends on AMR in Singapore and identify the types of research conducted. Scientific literature on AMR from Singapore published between 2009 and 2019 were retrieved from databases using a search string that included search terms that would encompass the range of terminologies related to “antimicrobial resistance” and “Singapore”. A total of 741 AMR research and review articles published between 2009 and 2019 were identified, which described research led by researchers from Singapore, that involved researchers from Singapore in overseas collaborations, or involved samples or data from Singapore. Articles were assigned to the most appropriate research domain and relevant sector(s) (animal, environment, food or human). Although an upward trend in the number of AMR research articles published was observed, articles that described research on AMR knowledge, awareness, socioeconomic impacts and transmission remained scarce. Furthermore, the higher proportion of research articles from the human sector highlighted that more research from the non-human sectors was needed, which coincidentally began to gradually increase in the last five years. By reviewing the types of studies that were conducted in each domain, broad areas where research gaps exist could be identified, as well as currently unexplored topics. With increasing complexity of the AMR problem and its impacts on multiple sectors, having a comprehensive overview of the evidence gaps is paramount to the development of a relevant One Health research agenda in AMR.

**Highlights:**

- A review of the trend and scope of AMR research including all sectors was conducted
- AMR research in Singapore is on the upward trend
- AMR research in the non-human sectors remained low
- Research on socio-behavioural factors and transmission of AMR to be prioritised

## Introduction

Antimicrobial resistance is a natural phenomenon, occurring as microorganisms evolve and compete for survival in their ecological niche [1]. However, the discovery of antimicrobials used in the treatment of infections, particularly antibiotics, has increased selection and accelerated the rate at which microorganisms acquire resistance [2]. Despite revolutionising human healthcare management worldwide, its misuse has led to the current global antimicrobial resistance (AMR) situation [3]. Reviews in the last decade provided alarming projections on the estimated healthcare costs and impact on global health [4, 5].

Microorganisms acquire resistance to antimicrobials through different mechanisms, including mutations, to gain survival advantage in the presence of antimicrobials. These mechanisms are quick to appear when antimicrobials are used [6], and cause a continuous feedback loop that leads to higher usage of antimicrobials to treat infections caused by increasingly resistant microorganisms. Research and development for new antimicrobials are needed when the existing ones are no longer effective.

Research has also gradually shifted towards identifying other non-biological factors that contribute towards the development of resistance. For example, the use of antimicrobials in livestock and agriculture for disease prevention and growth promotion purposes contributes to the increasing burden of AMR. Furthermore, antimicrobials are also used in companion animals, and the interaction between humans and companion animals can facilitate the spread of AMR microorganisms [7]. Therefore, multidisciplinary research that adopts a One Health approach [8] is needed to uncover the increasingly complex nature of AMR.

Scientific literature on AMR had been reviewed by others for specific microorganisms [9] or in specific sectors [10, 11] to study the global AMR research trends to guide future research directions for the global research community. For Singapore, such a review could help with understanding and proposing research needed at the local level to address AMR in the country, as well as to guide funding allocation in addition to charting future research direction at the national level. Therefore, we studied the scientific literature on AMR that had been published from Singapore between 2009 and 2019, with the aim of understanding the types of research performed to address AMR in Singapore, areas where research gaps still exist, and the local AMR research community. Articles that reported antimicrobial use (AMU) were also included as AMR and AMU are closely related.

## Methods

### Identification of Research Articles

To review the scientific literature on AMR in Singapore, we used the search string in Supplementary Information SI1 to retrieve peer-reviewed articles published on AMR between 2009 and 2019 from the following major bibliographic databases, without restrictions on language or study design to maximise search results: Global Health (Ovid), Scopus, Embase (Ovid), MEDLINE (Ovid), CINAHL (EBSCO), PubMed and Web of Science. We also manually checked the reference lists of included studies for potentially relevant articles.

### Study Selection

The search results from different databases were combined in a single EndNote library to remove duplicated records. A preliminary screen of all titles and abstracts was performed. Full texts of articles were then retrieved and read in full by SP and DMG, and included based on the eligibility criteria described below.

### Eligibility Criteria

Articles were included only when the main scope of research or review was on addressing AMR. Additional criteria included, but were not limited to, identifying methods to reduce or prevent the occurrence, prevalence or incidence of AMR; treating infections due to AMR microorganisms or developing methods to remove or improve removal of antimicrobials; and understanding the levels of antimicrobial use/AMR and its impact. We defined articles as being from Singapore (SGR) when it was published by authors from Singapore, identified by their institutions. Articles were defined as research collaborations when authors from Singapore were part of a study led by a research team from an overseas institution (SGO). Articles that used AMR samples or data collected from Singapore were also included (SGS).

We excluded studies that were pre-prints or e-published outside of the stipulated time frame; editorials, conference abstracts or proceedings, opinion pieces and guidelines; research that was not conducted in Singapore or by researchers from Singapore; as well as studies that did not have AMR as its main focus.

### Data Extraction, Analyses and Article Classification

Each article was reviewed manually to extract the study aim and outcomes, publication year, journal title, lead institution(s), microorganisms studied (to the genus level), and recorded in a standard form. Institutional information was determined by first and corresponding authors’ affiliations, as we hypothesised that they played a larger role in the conceptualisation of the article’s research focus [9], and used to determine whether the articles published should be categorised as SGR, SGO or SGS. Articles were read in full and assigned to the appropriate sector(s) (animal, environment, food or human) and one of nine research domains. The research domains were identified via a consultation with stakeholders in the field of AMR in Singapore (Table 1). When an article spanned multiple research domains, it was assigned by consensus to the most relevant single domain.

**Table 1.**
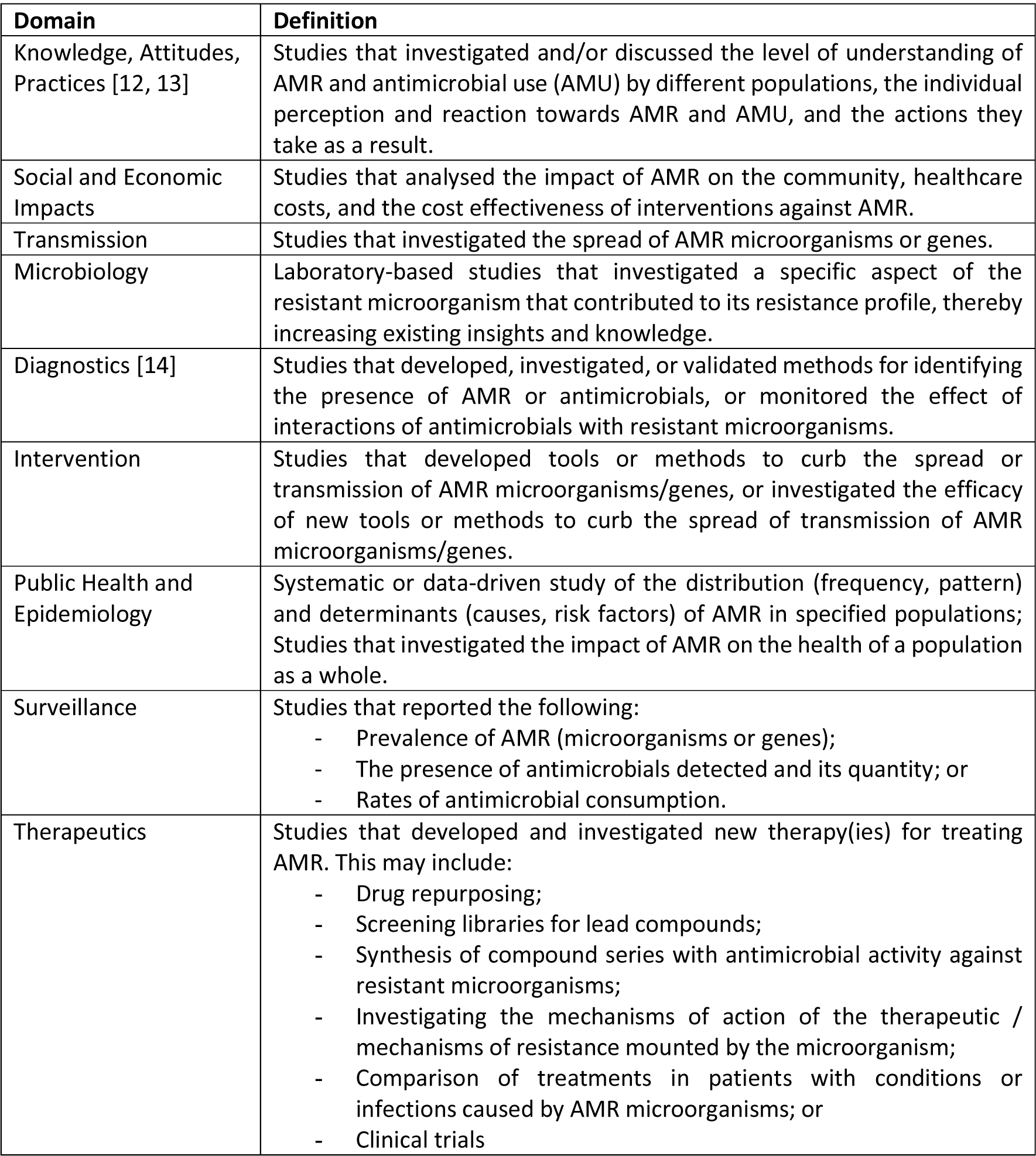
Research domains and definitions.

## Results

### Search results

Of 8700 articles from our initial search, we identified 717 eligible articles (Figure 1). Another 24 articles were identified through a manual search of references. In total, 741 articles were found to be relevant. Of these, 551 were SGR articles, 62 were SGS articles, and the remaining 128 were SGO articles (Supplementary Tables ST1 and ST2).

**Figure 1.**
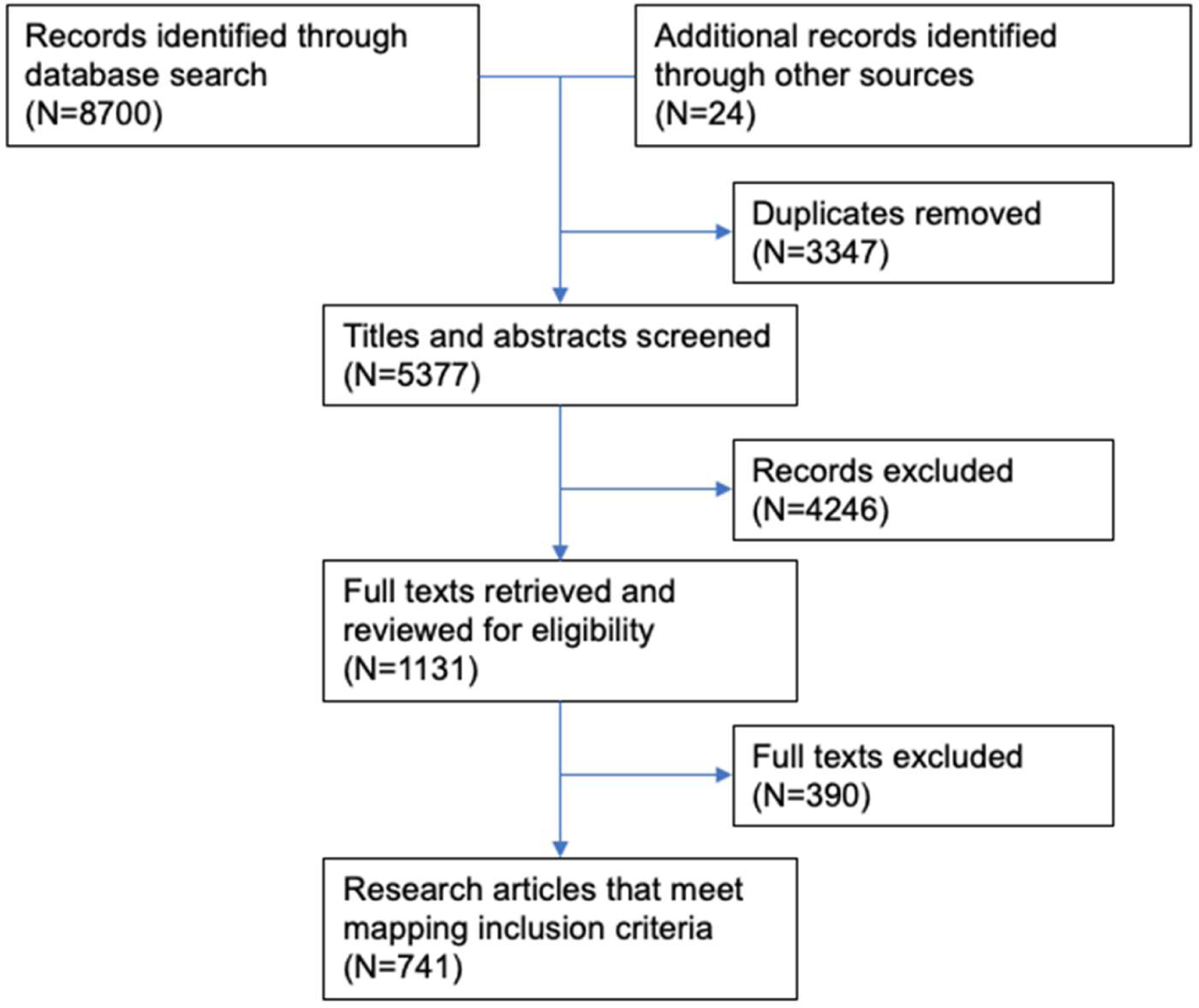
Flow diagram of article identification process.

### Trend of AMR research published from Singapore, 2009-2019

The number of SGR articles increased from 17 in 2009 to 73 in 2019, representing a threefold increase (Figure 2A). Most of the articles were from the human sector (N=490, 89%), followed by the environment (N=27, 5%) and food (N=10, 2%) sectors. Most of the research that covered more than one sector were from the human-environment sectors (N=14, 3%) (Figure 2B(i)). The annual trend of articles published from each sector also showed an increase between 2009 and 2019, with research articles from the environment and food sectors showing a prominent increase from 2017 (Figure 2Bii). Research on AMR from Singapore could be distributed to nine domains (Figure 2C(i)). The annual trend of research published from most domains was relatively consistent, except for the three domains with the lowest number of articles published: *Knowledge, Attitudes, Practices*, *Social and Economic Impacts* and *Transmission* (Figure 2Cii). Majority of the AMR research in Singapore was conducted by the institutes of higher learning (universities and polytechnics, N=245) and public healthcare institutions (N=260) (Supplementary Table ST3).

**Figure 2.**
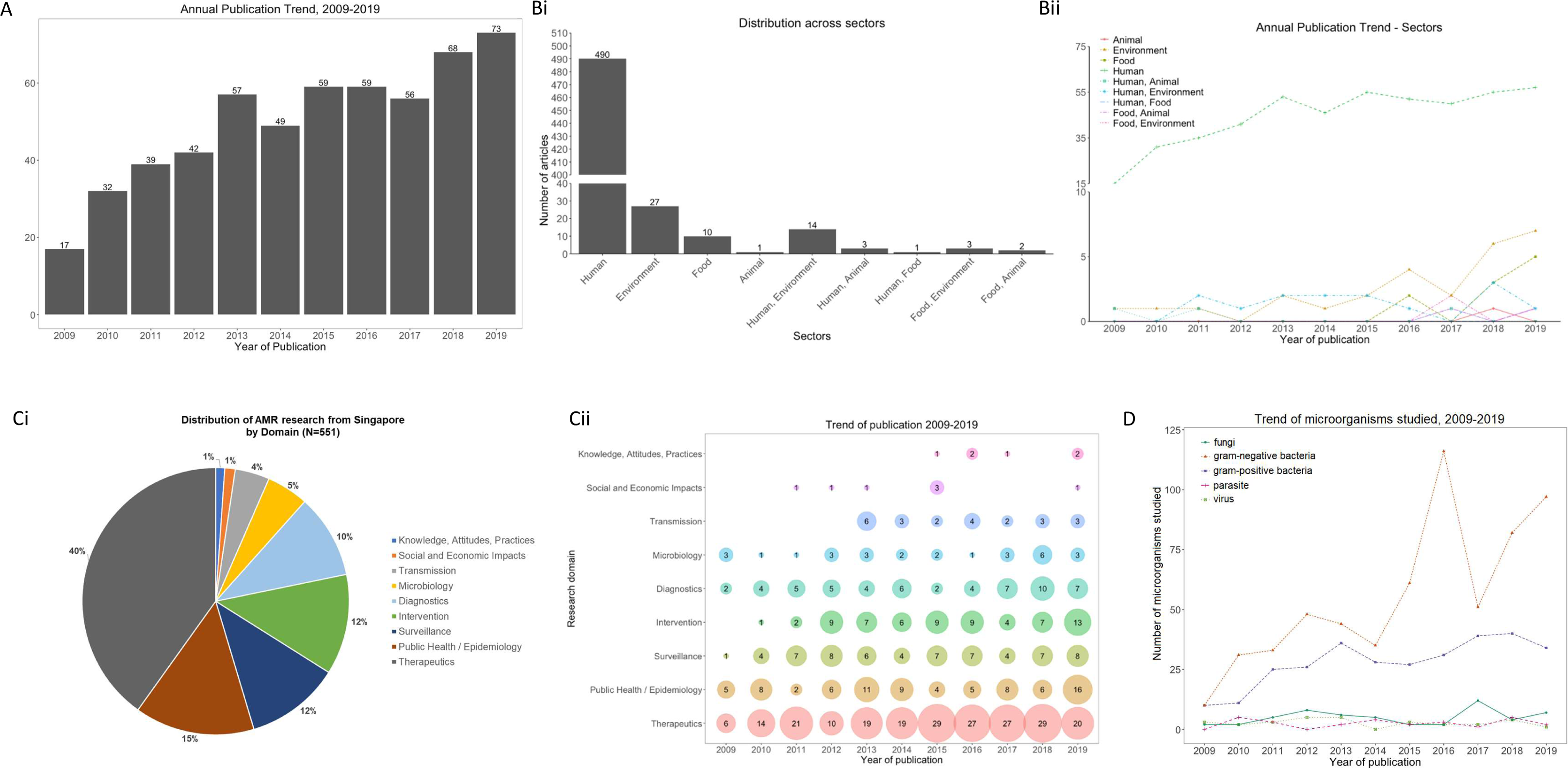
AMR research articles from Singapore published 2009-2019. (A) Number of articles published per year, from 2009 to 2019. (B) Number of research articles published in the One Health sectors (i) in total, and (ii) annually. (C) Distribution of research articles from Singapore across domains (i) in total, and (ii) annually. (D) AMR microorganisms studied across the years.

Most of the articles identified studied AMR in bacteria, both in terms of phenotypic and genotypic characterisation; AMR in gram-negative bacteria was studied more than in gram-positive bacteria (total 608 versus 307). In comparison, AMR in fungi, parasites or viruses was not studied as much but the frequency remained relatively consistent between 2009 and 2019 (Figure 2D). There was also active research conducted to identify or develop novel agents against AMR bacteria listed on the World Health Organization’s (WHO) priority pathogens list for therapeutics development [15] (Table 2).

**Table 2.** Number of research articles from the *Therapeutics* domain that focused on developing novel treatments for AMR pathogens on WHO’s priority pathogens list.

| AMR Pathogen | Number of articles |
| --- | --- |
| Priority 1: Critical |  |
| *Acinetobacter baumannii* | 11 |
| *Pseudomonas aeruginosa* | 43 |
| Enterobacteriaceae (Enterobacterales) | 40 |
| Priority 2: High |  |
| *Enterococcus faecium* | 9 |
| *Staphylococcus aureus*, methicillin-resistant | 59 |
| *Helicobacter pylori* | 2 |
| *Campylobacter* spp. | 0 |
| Salmonellae | 2 |
| *Neisseria gonorrhoeae* | 0 |
| Priority 3: Medium |  |
| *Streptococcus pneumoniae* | 1 |
| *Haemophilus influenzae* | 0 |
| *Shigella* spp. | 0 |

The types of studies in each domain were diverse, highlighting that understanding AMR required a multidisciplinary approach. Through reviewing the outcomes of AMR research from each domain that were already conducted, existing gaps were identified that could be potential areas for future research (Table 3 and Supplementary Information SI2).

**Table 3.** Summary of AMR research from Singapore in each research domain.

| Domain (N) | Sector(s)* (N) | Description of research | Summary of outcomes | Gaps |
| --- | --- | --- | --- | --- |
| Knowledge, Attitudes, Practices (6) | H (6) | <u>Healthcare professionals</u><br>Understand the medical and psychosocial determinants of antibiotic prescribing by physicians, and the factors influencing physicians' acceptance of recommendations provided by a computer decision support system for antibiotics prescribing.<br><u>Non-healthcare professionals</u><br>Investigate knowledge of appropriate antimicrobial use. | <u>Healthcare professionals</u><br>Factors associated with low or appropriate antibiotic prescribing practices were: good medical knowledge, clinical competency, good clinical practice and availability of diagnostic tests.<br>Lower threshold of prescribing antibiotics observed in emergency department physicians when treating patients who were old or had comorbidities.<br>Factors associated with acceptance of computer decision support system recommendations were related to seniority, experience and patient status.<br><u>Non-healthcare professionals</u><br>Overall knowledge of antibiotic use in the general population was poor.<br>Factors associated with higher antibiotic use were: ethnicity, housing type, high social economic status and lower education levels. | Effective education interventions to correct the misconception in the general population were needed.<br>Effective education modalities for each population that could subsequently be scaled up to target specific populations were needed. |
| Social and Economic Impacts (7) | H (7) | <u>Economic analyses</u><br>Studies analysed the expenses for treatment of infections caused by AMR microorganisms, cost effectiveness of intervention programmes (such as screening and isolation), and the cost effectiveness of accepting recommendations made by hospital's antimicrobial stewardship programme for treating patients with antibiotics.<br><u>Social and psychological</u><br>Studies analysed the social and psychological impact of infections caused by AMR microorganisms on patients. | <u>Economic analyses</u><br>Median cost or excess cost to treat patients with infections caused by AMR microorganisms were higher and significant.<br>There was also an increase in the length of stay and higher post-discharge care costs incurred.<br>Screen- and-isolate programmes were cost-effective and could reduce the rate of MRSA acquisition.<br>Recommendations made by hospital's antimicrobial stewardship programme reduced length of stay and costs incurred due to direct savings from reduced antibiotic use and subsequent readmissions.<br><u>Social and psychological</u><br>Rate of mortality was higher.<br>Patients experienced emotional trauma, depression, anxiety due to being isolated following a positive screening result. | Common limitation observed was that studies were limited by sample size or site, which limited subgroup analyses for impact identification.<br>Improvements in communication between healthcare workers and patients could be made to mitigate the psychological impact of isolation. |
| Transmission (23) | H (16)<br>H/A (1) | All studies investigated the transmission of AMR bacteria in the following situations: | <u>Investigation of outbreaks</u> | Nosocomial transmission: Patient care and handling |
|  | H/E (6) | • <u>Outbreaks of infection</u> caused by AMR bacteria in the intensive care unit or ward environment, in the population or community, and an animal experimental centre were investigated.<br>• Spread and dissemination of <u>antimicrobial resistance genes (ARGs)</u> found on plasmids, or the structure of transposon elements implicated in the spread of ARGs were investigated using gene sequencing as a common methodology.<br>• Role of <u>international travel</u> in MRSA transmission was discussed in a review. | • Outbreaks had an environmental source (built environment), or could be due to nosocomial, vector-borne or zoonotic transmission.<br>• Improving the built environment (by removing grooves in wash sinks) reduced transmission due to the environmental source.<br>• In the community, MRSA and TB transmission occurred at places where prolonged contact could occur, such as at gaming centres, in the prison and in the school.<br>• Sequencing methods identified the clonal dissemination of <u>ARGs</u>.<br>• Phage transduction, plasmid transconjugation, plasmid acquisition and mobile genetic elements were identified through sequencing as possible mechanisms of the spread of ARGs.<br>• <u>International travel</u> could play a significant role in transmission or introducing circulating MRSA strains to a country, with potential to replace existing endemic MRSA strains. | could be improved to limit nosocomial transmission. Studies on the types of improvements needed.<br>• Zoonotic: More studies were needed to determine whether zoonotic sources of AMR bacteria were endemic in the animal population and the potential impacts in confirmed cases of zoonotic transmission.<br>• International travel: Approaches to reduce introduction of AMR microorganisms from international travellers could be introduced to prevent onward transmission, and studies could be conducted to identify the effective approaches. |
| Diagnostics (56) | H (52)<br>E (3)<br>H/A (1) | Studies described the development of different approaches to detect or monitor AMR and antimicrobials. These methods could be segregated into:<br>• Assay-based<br>• Genetic approaches<br>• Imaging<br>• Spectrometry<br>• Predictive scoring<br>• Sample collection | <u>Assay-based</u><br>• Bioluminescence and colorimetric assay methods provided rapid turnaround time with accuracy.<br>• The reliability of commercial kits may be questionable for the purpose of detecting carbapenemases from different classes, but chromID CARBA SMART improved detection of OXA-CPE.<br>• Disk-based inhibitor tests worked better than agar dilution methods to determine drug susceptibility or AMR bacteria.<br><u>Genetic approaches</u><br>• Real-time, multiplex PCR, high-resolution melting curve analyses, sequencing and nucleic acid hybridisation were novel methods developed to correctly identify resistance mutations. | <u>Assay-based</u><br>• Host organ function was not a good proxy to detect antiviral drug resistance emergence for Hepatitis B virus infection.<br>• The best methods for measuring antibiotic synergy for pan- or extremely-drug resistant *Acinetobacter baumannii* had yet to be developed.<br><u>Genetic approaches</u><br>• Limitations in genotyping to accurately predict NRTI activity in HIV-1 patients receiving protease-based |
|  |  |  | <u>Imaging</u><br>Fluorophore-tagged compounds that could identify unique signatures or markers in AMR pathogens were developed. This allowed subsequent visualisation via fluorescent microscopy.<br><u>Predictive scoring</u><br>A prediction algorithm formulated based on logistic regressions of retrospective cohort of patients was validated in a prospective cohort.<br><u>Sample collection</u><br>The combination of anatomical sites most useful to accurately identify MRSA infections in dermatology, HIV and patients with infectious diseases was identified. In each group, the inclusion of perianal or throat swabs increased the sensitivity of MRSA detection. | antiretroviral therapy as second-line therapy. |
| Intervention (67) | H (49)<br>E (9)<br>F (3)<br>E/F (2)<br>H/E (4) | Studies described the development of tools or methods to curb the spread or transmission of AMR pathogens or genes, or investigate their efficacy in the following areas:<br><u>Healthcare interventions</u><br>Through antimicrobial stewardship and infection prevention and control<br><u>Bioengineering</u><br>Through the development of biomaterials and compounds with antimicrobial activities | <u>Healthcare interventions</u><br>Recommendations from hospital's antimicrobial stewardship teams led to shorter duration of therapy, length of stay and cost savings, and the recommendations did not compromise patient safety.<br>Antiseptics could be used in combination with prophylactic antibiotics to prevent catheter-related infections, such usage led to high prevalence of antibiotic resistant staphylococci in a dialysis unit, with suspected reduced susceptibility to the antiseptic.<br>Studies to investigate hand hygiene programmes showed that implementing ward-level accountability led to sustained improvement in hand hygiene compliance rate.<br><u>Bioengineering</u><br>Biomaterials that had antimicrobial activity with novel mechanisms of action were developed. Their manufacturing process was also described in some studies. | <u>Healthcare interventions</u><br>Further research needed to determine the quantitative association between increased hand hygiene compliance and use of alcohol-based hand-rubs with MRSA reduction.<br>The role of improved hand hygiene compliance for MRSA control, independent of contact precautions, should be elucidated. |
|  |  | <u>Engineering</u><br>Through treatment of waste to removal ARGs, or through modification of surfaces to prevent biofilm formation<br><u>Food safety</u><br>Through the removal of microorganisms with potential to harbour AMR from food. | Biomaterials could be applied as catheter coatings, and act as quorum sensing inhibitors, disrupt bacterial growth and motion, or inactivated bacteria.<br><u>Engineering</u><br>Constructed wetlands, membrane bioreactor or photoreactor systems in wastewater treatment processes were effective in removing ARGs.<br>Pre-treatment and addition of activated carbon enhanced ARG removal during solid waste treatment.<br>Surfaces modified with titanium dioxide or by physical texturing could reduce contamination by MRSA.<br><u>Food safety</u><br>The formation of biofilms in hydroponic farms or in food processing factories could be reduced by UV treatment or using sanitisers with quaternary ammonium compounds.<br>Novel plant extracts could provide an alternative to reduce or inhibit MRSA growth in food. |  |
| Public Health and Epidemiology (80) | H (78)<br>E (1)<br>H/A (1) | Studies in this domain focused on the following:<br><u>Modelling</u> the impact of antimicrobial selection pressure or consumption on the emergence of AMR.<br><u>Case reports</u> highlighting emerging or unique cases of infection caused by AMR pathogens.<br><u>Longitudinal observations</u> of AMR microorganism and spread.<br><u>Risk assessments</u> associated with AMR pathogens.<br><u>Qualitative study</u> of policy recommendations to address AMR. | A summary of the outcomes from the respective types of studies as follows:<br><u>Models</u> provide a tool to identify risk factors with highest impact on AMR and guide the selection of interventions. The models also provide a means to study possible response by microbial populations to the antimicrobials used.<br><u>Case reports</u> included literature reviews to highlight potential risks and recommendations to physicians when emerging or unique cases of infections caused by AMR pathogens were identified.<br><u>Longitudinal observations</u> provided an update on the susceptibilities of clinically relevant anaerobes causing bloodstream infections, while phylogenomic analyses on longitudinal samples provided an understanding on the genetic changes associated with adaptation in global spread of MRSA.<br><u>Risk assessments</u> provided and evaluation on the risk of importing resistant infections due to international travel and the mitigating measures that were already in place to counter the corresponding risks.<br><u>Qualitative studies</u> utilised semi-structured interviews with participants with relevant expertise to identify major | <u>Policies on AMR</u>: understanding how the social, political, cultural |
|  |  | • <u>Prevalence and distribution</u> of AMR microorganisms.<br>• Correlation of <u>microbial genotypic or phenotypic trait</u> with AMR.<br>• Correlation of <u>antimicrobial use with antimicrobial resistance</u>.<br>• Correlation of <u>risk factors, impact of or treatment outcomes</u> related to infection by AMR microorganisms. | themes on the areas of research that were needed. Insights on the social, political, cultural and behavioural aspects were also obtained.<br>• The <u>prevalence and distribution</u> of AMR microorganisms of genes conferring resistance were analysed. Results from the analyses provided information on the status of the resistance and the recommendations for monitoring potential risks of spread.<br>• <u>Genetic polymorphisms or mutations</u> identified in resistant *Candida auris* and *Mycobacterium tuberculosis* isolates correlated with changes in minimum inhibitory concentration to specific antimicrobials, were not present in susceptible isolates. Mutation conferring macrolide resistance in *Mycoplasma pneumoniae* could also be identified.<br>• Cross-correlation demonstrated possible associations between <u>the increased prescription of antibiotics to specific resistance</u> in *Escherichia coli* and *Acinetobacter spp.* Similarly, correlation was observed in chlorhexidine use and MRSA resistance. While oseltamivir-resistance in pandemic influenza A/H1N1/2009 was low, its observation emerged following the increased use of oseltamivir, prompting that prudent use and continued monitoring to be recommended<br>• With respect to <u>infection by AMR microorganisms</u>, the following were observed:<br>○ Mortality positively correlated with presence of resistance genes.<br>○ Colonisation by or carriage of AMR microorganisms positively correlated with longer length of stay at healthcare facilities, hospitalisation or outpatient visits, exposure to antimicrobials, medical devices or previous colonisation or carriage. | and behavioural aspects interact and shape the response by the population towards AMR. An understanding would aid in designing appropriate interventions to improve the population's response<br>• As an intervention to reduce infection by AMR microorganisms, <u>vaccination</u> should be recommended where available to reduce mortality (e.g. pneumococcal vaccination). |
| Surveillance<br>(63) | H (43)<br>E (10)<br>F (4)<br>A (1)<br>H/E (1)<br>H/F (1)<br>A/F (2)<br>F/E (1) | Several types of surveillance studies were identified and focused on the following:<br>Antimicrobial use<br>Prevalence and distribution of AMR microorganisms or genes<br>Molecular characterisation of emerging cases<br>Sources of data included:<br>Antimicrobial sales data<br>Existing surveillance programmes<br>Retrospective data<br>Point prevalence studies | Respective surveillance study types found the following:<br>Antimicrobial use in nursing homes in Singapore was generally low compared to other countries, including developed countries. However, the high percentage of inappropriate carbapenem prescription, and high rates of some antimicrobial agents suggest opportunities for antimicrobial stewardship.<br>Retrospective patient records reported prevalence of AMR microorganisms or genes, while data from the food and animal sectors identified resistant bacteria with potential for transmission. Abundance of antimicrobials and ARGs from the environment were reported, but longitudinal observation would be needed to monitor their trends.<br>Emerging cases of AMR microorganisms were reported, highlighting the importance of continued surveillance. | Using data from <u>environmental prevalence</u>, risk of ARGs being transferred into the aquatic environment could be assessed. |
| Therapeutics<br>(221) | H (219)<br>H/E (2) | The research could be further segregated into the following sub-domains:<br><u>Basic science</u><br>New compounds that exhibited effective antimicrobial activity against AMR microorganisms were identified, developed, synthesised or optimised. *In vitro* drug combinations, drug repurposing or screening of existing drug libraries were also used to identify new treatments.<br>Promising targets were further investigated in preclinical studies.<br>For some novel compounds, mechanisms of action were further investigated.<br><u>Pharmacokinetics-pharmacodynamics</u><br>Modelling using clinical data.<br><u>Biofilms</u><br>Identify and develop therapeutics that could be used to eradicate biofilms. | <u>Basic science</u><br>Novel materials or compounds generally demonstrated improved activity over existing antimicrobials against multidrug resistant organisms.<br>Besides showing good *in vivo* efficacy in reducing bacterial burden or parasitaemia, novel compounds also showed low cytotoxicity.<br>Hits from existing drug libraries with good efficacy and potential to be repurposed could be quickly approved for use as some of these compounds were already on the market.<br><u>Pharmacokinetic-pharmacodynamics (PK-PD)</u><br>Modelling provided optimal selection of empiric carbapenem dosing that was also cost effective, and achieved optimal cumulative fraction response in gram-negative infections.<br><u>Biofilms</u><br>Biofilms display higher resistance to antimicrobials. Studies identified novel compounds that, when used in combination with antimicrobials, could disrupt and inhibit | <u>Basic science</u><br>Novel compounds or materials with good *in vivo* efficacy from preclinical trials should be further developed for clinical trial testing.<br><u>PK-PD</u><br>Safety and efficacy of optimal doses modelled from retrospective data should be confirmed in the clinical setting. |
|  |  | Mechanism of biofilm resistance towards therapeutics was also investigated.<br><u>Bioinformatics</u><br>Mechanisms of action or resistance<br>Identify novel resistance targets<br>Identify potential new targets<br><u>Treatment</u><br>Retrospective analyses or case reports of treating infections caused by multidrug resistant microorganisms, clinical trials or the emergence of resistance following short duration of antimicrobial therapy. | the growth of biofilms, without using high concentrations of the antimicrobial.<br>Pulse-labelling or genetic methods were used to compare the differences in protein or gene expression between planktonic and biofilm bacteria. The differential expression of specific proteins or genes provided an explanation for the increased tolerance against antimicrobials.<br><u>Bioinformatics</u><br>*In silico* methods used to model the interaction between antimicrobials and microorganisms to elucidate how therapeutics work or how microorganisms evade the activity.<br>In viruses, structural analyses of critical viral proteins were performed to identify sites with lower threshold for resistance mutations and identify novel target sites for drug development that would have higher threshold to resistance development.<br><u>Treatment</u><br>Treatment outcomes reported were specific to the condition being treated.<br>Culture-guided step-down use of effective narrow-spectrum therapy, or using individualised combinations based on *in vitro* combination testing, were recommended for managing resistant bacterial infections.<br>Empirical therapy was recommended to be guided by pathogen susceptibilities to achieve desired optimal outcomes. | <u>Bioinformatics</u><br>Genetic mutations identified *in silico* to be putatively involved in drug resistance mechanisms could be further confirmed in *in vitro* and *in vivo* studies.<br><u>Treatment</u><br>Common limitations such as sample size, demographic representation of patients and the retrospective nature of the studies should be overcome with better study designs. |
| Microbiology (28) | H (20)<br>E (4)<br>F (3)<br>H/E (1) | Studies in this domain generally investigate a particular aspect of the microorganism to gain further understanding of its contribution towards resistance development. These include<br>The dynamics of <u>biofilm formation</u> in a co-culture of two bacteria species<br>Investigating the <u>genetic link to phenotypic resistance</u><br><u>Novel response to therapeutics</u> in bacteria or fungi<br>Identifying <u>novel mechanisms of resistance</u>. | Bacteria in co-culture could influence the dynamics of <u>biofilm formation</u>. Increase fitness of biofilms against antibiotics was observed.<br>The genetic-phenotypic links could be characterised in bacteria isolated from food, hospital wastewater and patients.<br>Chloroquine could induce programmed cell death pathway in chloroquine-resistant *Plasmodium falciparum*, providing novel targets for drug development. Response of *Candida* species to antifungals when serum was present could impact the minimum inhibitory concentration. | Microbiology studies provided opportunities to identify novel targets for drug development. |
|  |  |  | In isoniazid-resistant *Mycobacterium tuberculosis*, identification of novel mutations in addition to known mutations suggested that there are novel targets to be identified. |  |

\* Sector assignment was determined, where applicable, by the source of the isolate or sample, or by the research application.
A: Animal sector, which included companion animals and wildlife; E: Environment sector, which included the natural and built environment; F: Food sector, which included ready-to-eat food, food preparation facilities and animals reared for consumption; H: Human sector, which included clinical studies.

### Overseas research collaborations on AMR to further our understanding, 2009-2019

In parallel with the increasing number of AMR research and review articles from Singapore, there was an increasing trend in the number of overseas research collaborations in AMR. No specific trend was observed until 2014, when it began to show an increase (Figure 3, *“International collaborations”*). Most of the articles published were from the human sector (N=109, 85%), followed by environment (N=10, 7.8%) and food (N=2, 1.6%). There were six research articles that spanned multiple sectors (4.7%).

**Figure 3.**
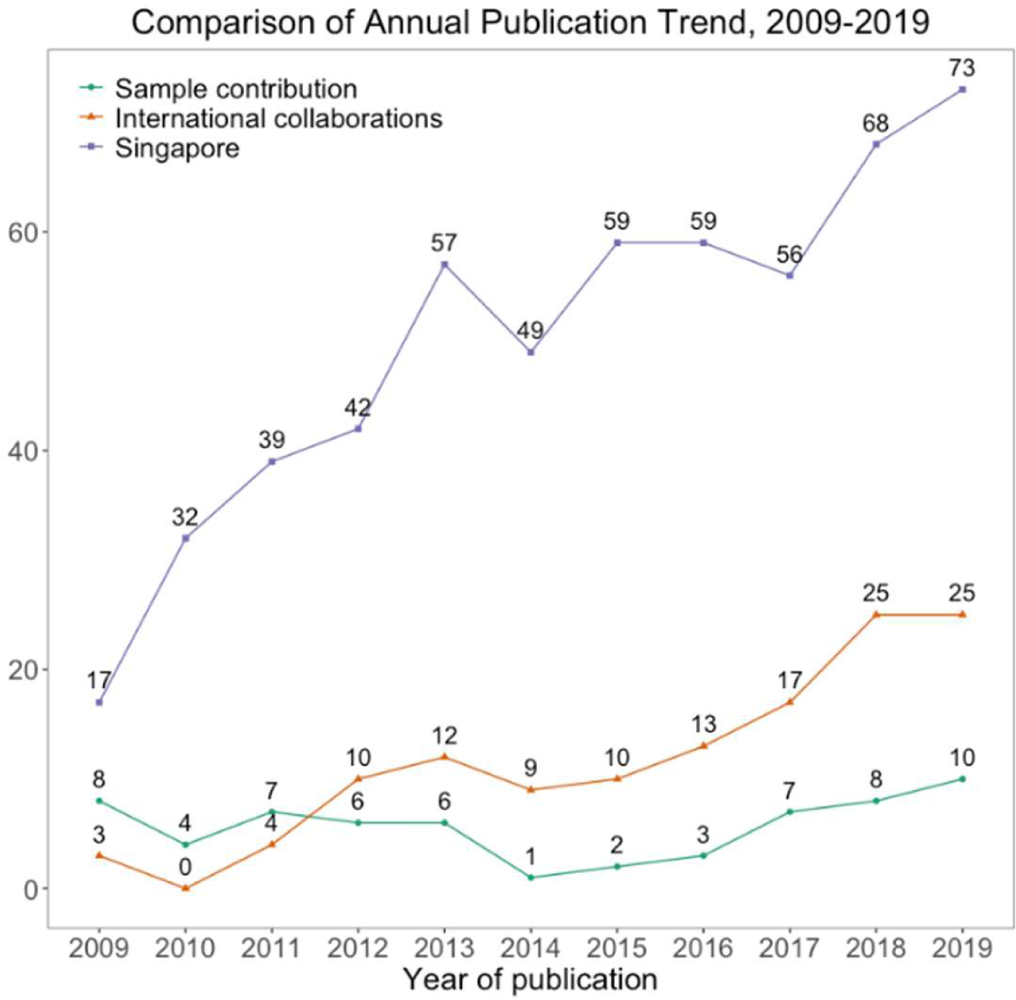
Comparison of AMR research trends from 2009 to 2019, for Singapore publications, international collaborations and research to which Singapore contributed samples.

The countries where the researchers had most collaborations with were Australia (N=24), followed by the United States (N=21) and the United Kingdom (N=14) (Figure 4A), with most research conducted on *Therapeutics* and *Diagnostics* (Supplementary Table 2). Within Asia, researchers from Singapore collaborated most with those from China (N=12), with the majority of the research conducted on *Surveillance*. Overall, the distribution of collaborative AMR research domains was similar to that of AMR research from Singapore, i.e. *Therapeutics* had the highest number of research articles, while *Knowledge, Attitudes, Practices* and *Social and Economic Impact* had the lowest number of research articles published (Figure 4B).

**Figure 4.**
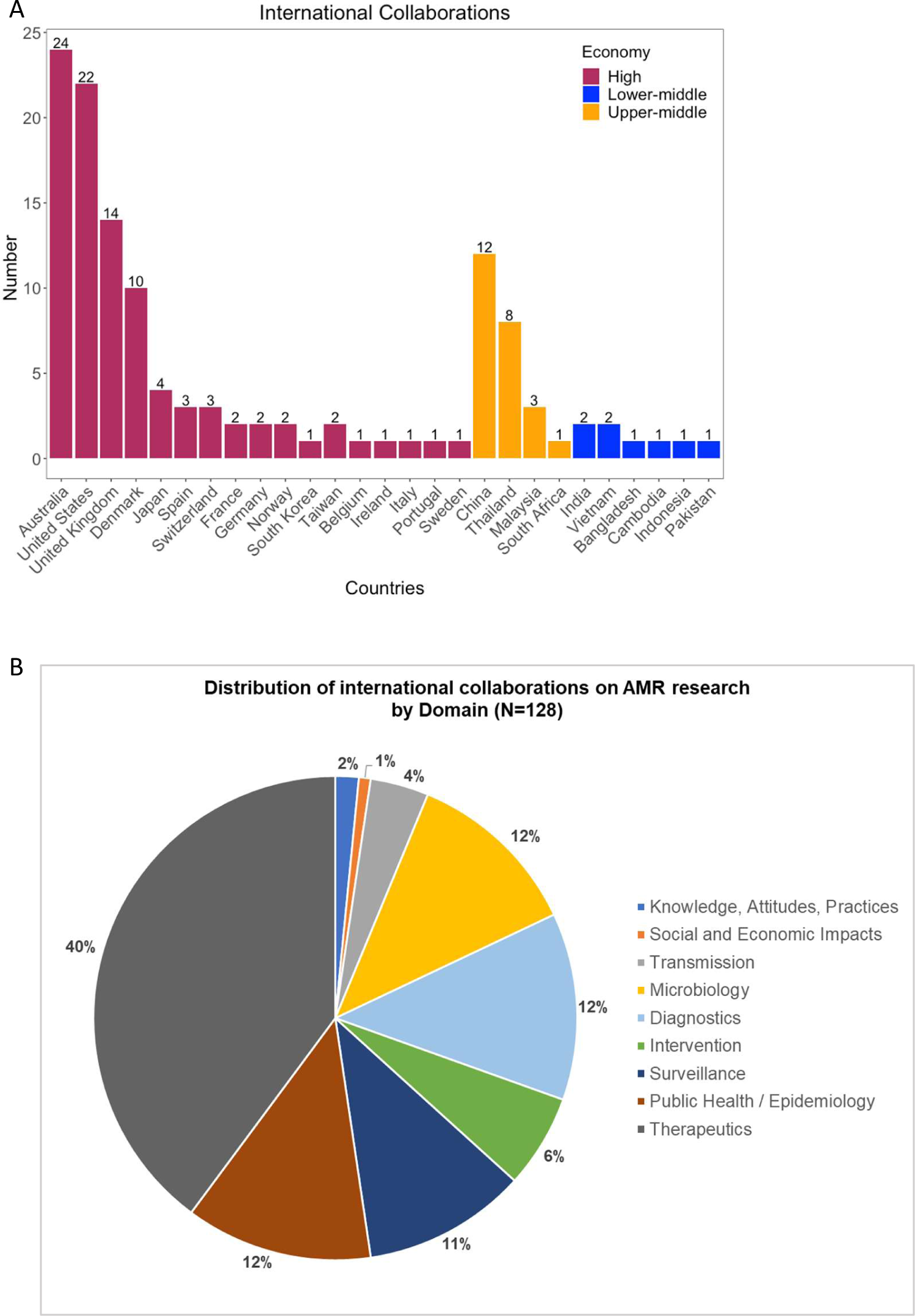
AMR international research collabroations, 2009-2019. (A) Number of AMR research articles, stratified by high, upper-middle and lower-middle income countries. Countries of origin are based on the institutional address of the lead or corresponding author. (B) Research trend by domain.

### AMR research that included samples from Singapore, 2009-2019

As part of ongoing multinational efforts to address AMR, researchers in Singapore participated in regional and global studies by contributing samples or data, or was a study site (Figure 3, *“Sample contribution”*). Of the 62 studies, 57 (92%) were from the human sector and four (7%) from the environment sector. One study was conducted across the human and animal sectors. These studies were assigned to the following research domains: *Surveillance* (N=39, 63%), *Public Health/Epidemiology* (N=16, 26%), *Therapeutics* (N=2, 3%), *Diagnostics* (N=2, 3%), *Transmission* (N=2, 3%) and *Microbiology* (N=1, 2%) (Supplementary Table 2). A description of the types of studies are listed in the Supplementary Table ST4.

## Discussion

The main objective of this article was to understand the AMR research conducted in Singapore in order to identify research gaps to guide future research. In addition, we also identified AMR research conducted internationally where Singapore collaborated in or contributed samples. Our data echoed other reviews of AMR research conducted in the wildlife [10], the food-producing animals [11], or the global AMR research [9], where an increasing trend was observed over time despite selecting different time periods for the review. For Singapore in particular, the launch of the National Strategic Action Plan (NSAP) on AMR in 2017 [16] coincided with a larger annual increase in the number of AMR research articles published. Furthermore, the quick succession of articles published on AMR research trends also indicated an interest to identify where research gaps exist to fill the gaps in our understanding.

However, unlike both reviews by Torres et al. [10, 11], our review did not limit the studies to a specific sector, or retrieve articles from a single database, as Luz et al. did [9]. The lack of such restrictions was to reduce as much bias as possible during the article retrieval stage, so that articles from all sectors or research fields could be retrieved as long as AMR was the main focus. By removing this bias, we could identify articles from all sectors, which highlighted the higher volume of articles from the human sector, in comparison to the non-human sectors. A quick comparison of research from single sectors alone showed that the combined number of articles from the animal, environment and food sectors was only 7.8% of the number of articles from the human sector. However, despite the small number, a gradual increase was observed after the launch of Singapore’s NSAP in 2017. Taking reference from the global observation and trend in AMR research from the wildlife and food-producing animals [10, 11], AMR research from the animal and food sectors in Singapore would likely continue in a similar trend. Furthermore, the increasing emphasis on understanding cross-sector influences and impacts of AMR [17], and the increasing evidence that antimicrobial misuse in food-producing animals harbours potential impact on human health [18] would also spur continued research from these sectors.

The three domains with the lowest number of research articles were *Knowledge, Attitudes, Practices*, *Social and Economic Impact*, *and Transmission*. Improving awareness and knowledge about AMR was a recurring theme regardless of geography or sector. In Japan, AMR awareness had not significantly changed despite WHO World Antimicrobial Awareness Week campaigns since 2015 [19]. Surveys conducted in Japan also showed continued misconceptions about antibiotics use [20], which was again echoed by a Chinese survey showing insufficient knowledge and neutral attitudes on AMR among consumers in the food sector [21]. Although culturally dissimilar, the similarities with a recent study on AMR knowledge and awareness among Singaporeans [22] demonstrated the need to improve AMR education. In addition to studies in Japan, China and Singapore, results regarding AMR knowledge in Ethiopia, Italy and Greece also showed that education with more appropriate content remained a requirement, and that this was not restricted to a single population or country [12, 23, 24]. Reviews on the contribution of AMR and the consequences for the population, such as that published by Verraes et al., provide updated information to the scientific community working on AMR in the food sector [25]. However, the information may not be easily understood by the public. Findings from a WHO survey on antibiotic awareness campaigns held globally recommended dedicating sufficient resources to campaign development and implementation, especially with updating obsolete information and using more effective communication strategies [26]. Campaign evaluations were also needed to measure their effectiveness and impact, since less than half of known campaigns were evaluated and the results of such evaluations were usually not publicly available [26].

Our review also highlighted that research that delved deeper into the contribution of each sector towards the development of AMR was needed. The environment, for example, is a known reservoir of *de novo* resistance development, but awareness and understanding of its potential impact are lacking [27-29]. The presence of AMR genes and bacteria in the environment continued to be an area of concern, with several meetings held in the last five years to formulate and update the research agenda on water, sanitation and AMR [30, 31]. Treatment systems in Singapore are effective in removing most AMR genes and bacteria, but research in this area is still ongoing to further improve the current treatment efficiencies [32-35]. Recommendations on priority research topics from global expert consultations such as these could also be adapted for Singapore’s context.

Singapore is exposed to AMR from multiple sources, such as from imported food or international travellers, and research on potential transmission pathways of AMR is important. Therefore, continued national, regional and global surveillance and epidemiological studies not only provide information on the prevalence and incidence of AMR in and around Singapore, but also update risk assessments and mitigation actions to minimise impacts where necessary [36-38]. Combining AMR risk assessments with potential social and economic impact assessment may provide a holistic approach towards overall impact assessment [39] and encourage the adoption of actions to minimise the impacts due to AMR. Research on the economic impact and burden due to AMR could be strengthened to facilitate such assessments for Singapore.

Lastly, the role of the animal, environment and food sectors in the transmission of AMR and their intersecting impact on all the One Health sectors should also be investigated [17]. Most of the current research conducted in Singapore addressed AMR from a human perspective, or from a single sector. AMR genes from the environment had been shown to impact human health, including extended-spectrum beta-lactamase and New Delhi metallo-beta-lactamase-1 genes [40, 41]. These two genes revealed a feedback loop between infection, antibiotic use and the development of antibiotic resistance [42]. Identifying and understanding the transmission of such genes from the environment, food and animal sectors would be necessary to plan risk assessments and mitigation measures. Being an island state where all sectors are in close proximity or sharing the same space, this information is important to minimise cross-sector transmission that would be detrimental to Singapore’s health security and economy.

Besides the three domains identified where research needs to be strengthened, other areas such as diagnostics for cross-sector application and an integrated surveillance system to facilitate correlation of data should also not be overlooked. With WHO working towards a global research agenda on AMR [43], Singapore could also identify relevant research questions needed to drive a national AMR research agenda that supports local needs using a similar approach.

### Strengths and Limitations

Like the three reviews of global research trends [9-11], this was the first review of Singapore’s research landscape on AMR. We did not limit the types of research in our search of articles from the databases, which allowed us to identify studies that could be assigned to the research domains identified from stakeholder consultation. The studies were also not limited in terms of research technique employed, and covered basic biological studies, sociology, public health, computational modelling and clinical trials. We also did not impose restrictions on language, although our initial search identified only articles published in English. However, this landscape review could have been limited by the lack of a key word analysis to further corroborate the domains identified. There was also a risk of excluding relevant research not indexed in scientific databases we searched, but this was mitigated by a manual check of the references of included articles. While posters presented at scientific meetings were excluded, this was unlikely to significantly impact the landscape review reported here as they represented ongoing research work that were subsequently published, and would likely still be included in our review. Due to the lack of agreed definitions for the sectors globally, articles were also categorised using self-defined sectors. Nevertheless, the review of Singapore’s AMR research landscape will be a continuous effort to identify the evolution in response to the global call and needs in AMR.

## Conclusion

This review provides the first overview of the AMR research in Singapore. Besides an increasing research output and diversifying topics in AMR research, it uncovered research gaps where resources could be dedicated to increase the amount of research and corresponding output needed to drive the AMR agenda highlighted by the Global Action Plan on AMR. In line with the One Health approach to AMR, more research on AMR in and between all sectors is encouraged to identify transmission between the One Health sectors. Collaborations with industry partners would also be needed to translate findings into practice. The population’s knowledge and response towards AMR should be improved, and research on effective education methods is needed. Furthermore, developing a better understanding of the direct and indirect costs of AMR would support the channelling of resources and optimisation of interventions. Innovative cross-disciplinary research should be encouraged and fostered, using a design theory approach that would break the current mould that researchers often limit themselves to [44].

## Author statement

SP and DMG were involved in all stages of the study; AK, THL, YNL and YSL reviewed and edited reviewed and edited the manuscript.

Parts of the review were presented at the 1^st^ International Symposium for Infectious Diseases Research Institutes Cooperation (22-23 Feb 2023).

## Data Availability

All data produced in the present work are contained in the manuscript.

## Acknowledgements

The research did not receive any specific grant from funding agencies in the public, commercial or not-for-profit sectors.

The authors would like to thank Ms Yasmin Lynda Munro from the Nanyang Technological University, Lee Kong Chian School of Medicine Library, for providing advice and assistance on our search of articles from the databases; Ms Hui Min NG, Ms Xiao Wei LIM and past interns and staff at National Centre of Infectious Diseases for their assistance with checking references, identifying and reading the research articles, and extraction of data.

Although the authors identified research articles published by clinicians, pharmacists and microbiologists from either Tan Tock Seng Hospital or National Centre for Infectious Diseases for this review, all articles were given equal treatment during the review process conducted for this publication.

