## Supplementary Information and Tables for "Antimicrobial resistance research in Singapore – mapping current trends and future perspectives"

### Supplementary Information SI1 – Search string for retrieval of articles from databases

The following was the search string for NCBI PubMed, and adapted accordingly for Embase (Ovid), Scopus, Cumulative Index to Nursing and Allied Health Literature (CINAHL, Ebsco), Global Health (Ovid), Medline (Ovid) and Web of Science.

| | |
|---|---|
| 1 | ((((((anti*) OR (anti-b*)) OR (anti-m*)) OR (anti-f*)) OR (anti-v*)) OR (microb*)) OR (antimicrob*)) OR (anti-microb*) |
| 2 | ((resistant) OR (resistance)) OR (resist*) |
| 3 | #1 OR #2 |
| 4 | (Singapore) OR (Singaporean) |
| 5 | #3 AND #4 |
| 6 | #5 AND ((“2009/01/01)”[Date – Publication] : “2019/12/31”[Date – Publication])) |

### Supplementary Information SI2 – Summary of AMR research from Singapore in each research domain

*Knowledge, Attitudes, Practices*

Articles identified in this domain involved studies that were conducted in the health sector, among healthcare professionals and non-medically trained populations. Among healthcare professionals, studies found that good medical knowledge, clinical competency, good clinical practice and availability of diagnostic tests were factors associated with low or appropriate antibiotic prescription [1, 2]. While there was overall willingness to consult with a computerised decision support system (CDSS) to aid in appropriate prescribing of antimicrobials, physicians' acceptance of CDSS recommendations was still related to their seniority, experience and patient status [3]. This was observed with experienced physicians who would tend to override recommendations when presented with a complex patient case with multiple infections or allergies. Similarly, physicians in the emergency department would prefer to prescribe antibiotics when faced with patients who were old or had comorbidities [2]. In non-healthcare professionals, the overall knowledge of antibiotic use was poor. Factors associated with higher antibiotic use were ethnicity, housing type, higher socioeconomic status and lower education [4-6].

*Social and Economic Impact*

The median or excess cost to treat infections due to AMR microorganisms were significantly higher, resulted in increased length of stay and post-discharge care costs [7-9]; the overall mortality rate was also higher. The implementation of interventions that prevented AMR transmission led to cost-savings, as evidenced by studies that analysed the cost-effectiveness of AMR interventions, such as screen and isolate programmes [10] and when recommendations from institutions' antimicrobial stewardship programmes (ASP) were adopted [11]. Screen and isolate programmes also led to reduction in MRSA acquisition rates [10], which had implications on the quality of healthcare delivery in hospitals [9]. However, the impact of screen and isolation programmes should be further studied as one study found that such isolation could cause emotional trauma, depression and anxiety [12]. Infections due to AMR also affected the dynamics between the patient and their social circle [13].

*Transmission*

An important scope of AMR research was to understand the transmission of antimicrobial resistant microorganisms and antimicrobial resistant genes (ARGs). Sources of transmission of AMR microorganisms could come from contamination within the built environment or surfaces [14-18]; the transmission would often be terminated when the root cause was removed. Studies also identified sources causing nosocomial [19-21], community [22-25], vector-borne [26] or zoonotic transmissions [27], with some recommendations of mitigation approaches; for example, to improve patient care and handling when healthcare staff were implicated as the source of MRSA transmission within healthcare facilities [19-21]. International travel could also impact on MRSA transmission by introducing emerging, circulating strains that could replace existing endemic MRSA strains. In this scenario, targeted screening of travellers at risk was recommended [28].

At the molecular level, phage transduction, plasmid transconjugation or acquisition, and the presence of mobile genetic elements were identified as possible contributors to increased transmission [29-36]. Insights from sequencing studies provided avenues for the development of appropriate intervention strategies to control their spread, one of which was the implementation of whole-genome sequencing as a possible infection prevention and control approach [37].

*Diagnostics*

Diagnostic tools are used to identify the presence of AMR pathogens or genes in infections for timely application of appropriate treatments, or for public health surveillance of the AMR. Research and development in diagnostics had been a continuous effort to improve accuracy, reliability and turnaround time of existing methods, or to validate new tools or methods. Majority were for application in the human sector and likely met the needs of healthcare professionals in diagnosing whether infections were caused by AMR pathogens. For instance, assay-based methods focused on either improving the accuracy and turnaround time to identify antibiotic combinations that were effective against carbapenem-resistant Gram-negative bacteria infections [38-40], or to ensure reliability of commercial tests [41, 42], and existing drug susceptibility methods [43-50]. Laboratories also developed genetic-based methods to identify the presence of antimicrobial resistance genes (ARGs) via polymerase chain reaction [51-59], high-resolution melting curve approach [60-63], or sequencing [64-68]. Unique markers of resistant pathogens allowed development of fluorophore-conjugated tags to visualise them via microscopy methods [69-73]. Three studies described the development of diagnostics for samples from the environment: these involved spectrometry and enabled identification of the presence of antimicrobials from water samples [74-76].

#### *Interventions*

Articles on intervention were focused on tools or methods to curb the development or spread of AMR pathogens. They included actions taken by infection control teams to prevent onward transmission at source in healthcare settings [77-94], or developing novel biomaterials that were effective in preventing growth of AMR pathogens or biofilms [95-97] that could be applied on medical equipment such as catheters [98-104]. Studies on antimicrobial stewardship recommendations were also included, which provided evidence that antibiotic use could be reduced without compromising treatment outcomes for the patient [105-118]. From the environment sector, waste treatment methods were developed and evaluated for their effectiveness in removing ARGs [119-123], while solid surfaces could be modified with nanoparticles or by physical texturing to prevent contamination or colonisation by resistant bacteria [97, 124, 125]. Interestingly, researchers were also looking into ways to curb AMR pathogen colonisation and growth in food [126-128].

#### *Public Health and Epidemiology*

Public health and epidemiology studies were closely related, as the findings from epidemiological studies, in terms of understanding prevalence, incidence, and underlying risk factors of AMR infection, could be applied to benefit the health of the population as a whole. Research articles assigned to this domain were also diverse in the types of studies carried out. Besides reporting on the prevalence and distribution of antimicrobial resistance rates, studies went on to analyse the underlying genetic associations [129-143] or mutations [144-146], antimicrobial use [147, 148], or with clinical condition [149-166].

Data collected in some studies also facilitated the identification of risk factors for acquisition of or colonization by resistant microorganisms [167-179], and model the impact of antimicrobial consumption on resistance development to guide treatment choices [180, 181]. Such data were also applied to risk assessments [182, 183]. Besides quantitative information, qualitative data or reviews holistically addressed the overall impact of antimicrobial resistance development on population health [184-192], with views on how antimicrobial resistance programmes could take steps to mitigate [193-195].

#### *Surveillance*

Surveillance studies in the human sector analysed retrospective patient records to report the prevalence of AMR microorganisms or related genes [196-213], the presence and quantity of antimicrobials detected, or antimicrobial usage [214-218]. Surveillance from the food [219-225] and animal [226] sectors identified resistant bacteria with potential to further spread AMR. Surveillance studies also analysed the abundance of antimicrobials (208) or resistance genes from the environment [226-234]. Beyond surveillance within a single sector, cross-sectoral surveillance had been valuable in revealing potential sources of transmission [235] and provided insights on resistance profiles of bacteria circulating amongst humans [236]. In addition to highlighting emerging cases of resistant microorganisms [237-255], data from surveillance had also indicated when outbreak investigations may be necessary due to the abnormal number of cases [221]. Surveillance efforts outside of Singapore also provided information on the resistance profiles present in different geographical regions that could inform healthcare professionals on the appropriate antibiotics to prescribe to mitigate further resistance development [209, 256-283]. These regular updates on resistance patterns lend insight to healthcare professionals on how patients should be managed in the era of global travel.

#### *Therapeutics*

The therapeutics domain consisted of research focused identifying effective treatments against drug-resistant microorganisms or biofilms. Basic science studies that identified, synthesized, engineered or modified promising compounds to improve their activities against resistance microorganisms were included; these studies often showed that the new compounds were more effective than existing antibiotics. Preclinical studies were also performed for some of these compounds [284-303]. Existing antibiotics were also investigated to identify possible combinations that could improve their minimum inhibitory concentrations (MIC) [304-307]. Approved drug libraries were also screened for potential repurposing [308-317]. In some studies, the mechanisms of action or resistance were studied, either using laboratory-based methods [318-345], or via *in silico* methods using bioinformatics tools that allow structural modelling [317, 346-354].

Where antimicrobials were already in use for treatment, studies were often retrospective analyses to report the benefits of using culture-guided step-down therapies for managing the specific condition or infection, or using information on pathogen susceptibilities to guide empirical treatments [355-380]. Pharmacokinetics-pharmacodynamics modelling was also performed [381]. However, such studies were often limited by the sample size and patient demographics, and called for the need for prospective cohort studies. Even with clinical trials were conducted [382, 383], results generally demonstrated that the treatment under investigation were non-inferior to existing treatments in general.

*Microbiology*

There were some research articles identified from this review that did not fit the definitions for the other domains, but had investigated specific characteristics of microorganisms to generate further insights on AMR. Therefore, these articles were assigned to the microbiology domain. These studies described within these articles included analysing the dynamics of biofilm formation in a co-culture of two bacteria species [384], correlating genetics to resistance phenotypes [385-391], studying the evolution of AMR development in bacteria [392-395], identifying novel responses or mutations to therapeutics in bacteria or fungi [396, 397], and investigating resistance mechanisms [398-407]. Knowledge gained from these studies could provide avenues for the identification of novel targets for therapeutics development.

### Supplementary Tables

#### Supplementary Table ST1. List of AMR research and review articles published by researchers from Singapore between 2009 and 2019.

Articles were assigned to the most relevant research domain and sector(s).

| Article ID | Sector(s) | Research Domain |
|---|---|---|
| 2009, Chan KS [238] | Human | Surveillance |
| 2009, Chuwa EWL [408] | Human | Public Health and Epidemiology |
| 2009, Deepak RN [132] | Human | Public Health and Epidemiology |
| 2009, Donaldson AD [409] | Human | Therapeutics |
| 2009, Fan C [410] | Environment | Microbiology |
| 2009, Fong RKC [411] | Human | Public Health and Epidemiology |
| 2009, Ho YM [400] | Human | Microbiology |
| 2009, Koh TH [142] | Human, Animal | Public Health and Epidemiology |
| 2009, Lee CC [177] | Human | Public Health and Epidemiology |
| 2009, Lim T-P [412] | Human | Therapeutics |
| 2009, Liu L [413] | Human | Therapeutics |
| 2009, Luan J [414] | Human | Diagnostics |
| 2009, Prabakaran M [357] | Human | Therapeutics |
| 2009, Tan TY [44] | Human | Diagnostics |
| 2009, Teo JWP [387] | Human | Microbiology |
| 2009, Tin S [415] | Human | Therapeutics |
| 2009, Wang S-Q [353] | Human | Therapeutics |
| 2010, Cheow WS [416] | Human | Therapeutics |
| 2010, Cheow WS [417] | Human | Therapeutics |
| 2010, Ch'ng J-H [397] | Human | Microbiology |
| 2010, Ding C [192] | Environment | Public Health and Epidemiology |
| 2010, Donaldson AD [204] | Human | Surveillance |
| 2010, Gan LSH [60] | Human | Diagnostics |
| 2010, Ho J [187] | Human | Public Health and Epidemiology |
| 2010, Hsu L-Y [147] | Human | Public Health and Epidemiology |
| 2010, Hsu L-Y [418] | Human | Therapeutics |
| 2010, Husain N [334] | Human | Therapeutics |
| 2010, Inoue M [419] | Human | Therapeutics |
| 2010, Jayaraman P [420] | Human | Therapeutics |
| 2010, Koh TH [137] | Human | Public Health and Epidemiology |
| 2010, Koh TH [242] | Human | Surveillance |
| 2010, Kurup A [83] | Human | Intervention |
| 2010, Lim LG [421] | Human | Diagnostics |
| 2010, Lim PL [422] | Human | Public Health and Epidemiology |
| 2010, Liu R [40] | Human | Diagnostics |
| 2010, Ng ES-T [423] | Human | Therapeutics |
| 2010, Nzila A [424] | Human | Therapeutics |
| 2010, Ong DCT [61] | Human | Diagnostics |
| 2010, Rottmann M [297] | Human | Therapeutics |
| 2010, Samy RP [298] | Human | Therapeutics |
| 2010, Soe WM [425] | Human | Therapeutics |
| 2010, Soe WM [426] | Human | Therapeutics |
| 2010, Sun Y-J [145] | Human | Public Health and Epidemiology |
| 2010, Tan TY [134] | Human | Public Health and Epidemiology |
| 2010, Tan TY [209] | Human | Surveillance |
| 2010, Vasoo S [212] | Human | Surveillance |
| 2010, Wijaya L [189] | Human | Public Health and Epidemiology |
| 2010, Wong L [427] | Human | Therapeutics |
| 2010, Zhou C [428] | Human | Therapeutics |
| 2011, Bahadin J [150] | Human | Public Health and Epidemiology |
| 2011, Chan HLE [237] | Human | Surveillance |
| 2011, Chantratita N [321] | Human | Therapeutics |

| Article ID | Sector(s) | Research Domain |
|---|---|---|
| 2011, Cheow WS [429] | Human | Therapeutics |
| 2011, Ch'ng J-H [320] | Human | Therapeutics |
| 2011, Fan C [394] | Environment | Microbiology |
| 2011, Hsu L-Y [205] | Human | Surveillance |
| 2011, Husain N [333] | Human | Therapeutics |
| 2011, Kyaw BM [430] | Human | Therapeutics |
| 2011, Kyaw BM [431] | Human | Therapeutics |
| 2011, Lee CY [432] | Human | Therapeutics |
| 2011, Lee HK [62] | Human | Diagnostics |
| 2011, Leung GYC [433] | Human | Therapeutics |
| 2011, Liew YX [216] | Human | Surveillance |
| 2011, Liew YX [434] | Human | Diagnostics |
| 2011, Liew Y-X [215] | Human | Surveillance |
| 2011, Lim PL [244] | Human | Surveillance |
| 2011, Lim T-P [307] | Human | Therapeutics |
| 2011, Lim T-P [306] | Human | Therapeutics |
| 2011, Lingegowda PB [435] | Human | Therapeutics |
| 2011, Mirza H [45] | Human, Animal | Diagnostics |
| 2011, Mok S [340] | Human | Therapeutics |
| 2011, My NH [436] | Human | Therapeutics |
| 2011, Nederberg F [437] | Human | Therapeutics |
| 2011, Ng LSY [82] | Human, Environment | Intervention |
| 2011, Ong DCT [56] | Human | Diagnostics |
| 2011, Pada SK [9] | Human | Social and Economic Impacts |
| 2011, Phua CK [370] | Human | Therapeutics |
| 2011, Saeidi N [438] | Human | Therapeutics |
| 2011, Samy RP [439] | Human | Therapeutics |
| 2011, Sim SH [440] | Human, Environment | Therapeutics |
| 2011, Soe WM [441] | Human | Therapeutics |
| 2011, Suhaila M [442] | Human | Therapeutics |
| 2011, Tan TY [46] | Human | Diagnostics |
| 2011, Teo BW [89] | Human | Intervention |
| 2011, Teo BW [165] | Human | Public Health and Epidemiology |
| 2011, Teo JWP [250] | Human | Surveillance |
| 2011, Vasoo S [213] | Human | Surveillance |
| 2011, Xing B [443] | Human | Therapeutics |
| 2012, Bai Y [319] | Human | Therapeutics |
| 2012, Balm MND [201] | Human | Surveillance |
| 2012, Cai Y [202] | Human | Surveillance |
| 2012, Chen CP [124] | Human, Environment | Intervention |
| 2012, Chen T [444] | Human | Intervention |
| 2012, Chen Y-T [239] | Human | Surveillance |
| 2012, Cheong CSJ [196] | Human | Surveillance |
| 2012, Chew KK [66] | Human | Diagnostics |
| 2012, Chien JMF [379] | Human | Therapeutics |
| 2012, Choudhury S [203] | Human | Surveillance |
| 2012, Choudhury S [50] | Human | Diagnostics |
| 2012, Chow A [445] | Human | Diagnostics |
| 2012, Chow A [79] | Human | Intervention |
| 2012, Cui Y [323] | Human | Therapeutics |
| 2012, Du H [446] | Human | Microbiology |
| 2012, Fukushima K [447] | Human | Therapeutics |
| 2012, Han N [330] | Human | Therapeutics |
| 2012, Huang Y [289] | Human | Therapeutics |
| 2012, Kanagarajan V [448] | Human | Therapeutics |
| 2012, Koh TH [241] | Human | Surveillance |
| 2012, Koh TH [140] | Human | Public Health and Epidemiology |
| 2012, Kyaw BM [449] | Human | Therapeutics |

| Article ID | Sector(s) | Research Domain |
|---|---|---|
| 2012, Win MK [152] | Human | Public Health and Epidemiology |
| 2012, Lee ASG [63] | Human | Diagnostics |
| 2012, Lee CK [65] | Human | Diagnostics |
| 2012, Liew YX [105] | Human | Intervention |
| 2012, Liew YX [114] | Human | Intervention |
| 2012, Ling ML [78] | Human | Intervention |
| 2012, Lye DC [157] | Human | Public Health and Epidemiology |
| 2012, Ng E [8] | Human | Social and Economic Impacts |
| 2012, Poon LM [151] | Human | Public Health and Epidemiology |
| 2012, Sarathy JP [407] | Human | Microbiology |
| 2012, Shao Q [450] | Human | Intervention |
| 2012, Teo JQM [171] | Human | Public Health and Epidemiology |
| 2012, Teo JQM [113] | Human | Intervention |
| 2012, Teo JWP [253] | Human | Surveillance |
| 2012, Vasoo S [175] | Human | Public Health and Epidemiology |
| 2012, Venkatachalam I [254] | Human | Surveillance |
| 2012, Verrall AJ [360] | Human | Therapeutics |
| 2012, Wozniak M [451] | Human | Therapeutics |
| 2012, Yeo C-L [109] | Human | Intervention |
| 2012, Zou G [405] | Human | Microbiology |
| 2013, Ang TL [368] | Human | Therapeutics |
| 2013, Balm MND [200] | Human | Surveillance |
| 2013, Balm MND [174] | Human | Public Health and Epidemiology |
| 2013, Balm MND [141] | Human | Public Health and Epidemiology |
| 2013, Balm MND [18] | Human | Transmission |
| 2013, Chee CBE [24] | Human | Transmission |
| 2013, Chin W [452] | Human | Therapeutics |
| 2013, Ch'ng J-H [309] | Human | Therapeutics |
| 2013, Chong C-W [453] | Human | Public Health and Epidemiology |
| 2013, Chow WL [125] | Environment | Intervention |
| 2013, Chua SL [454] | Human | Therapeutics |
| 2013, Fisher D [92] | Human | Intervention |
| 2013, Fukushima K [287] | Human | Therapeutics |
| 2013, Goh S [35] | Human | Transmission |
| 2013, Grant D [22] | Human | Transmission |
| 2013, Heng YK [455] | Human | Therapeutics |
| 2013, Hon PY [54] | Human | Diagnostics |
| 2013, Koh J-J [335] | Human | Therapeutics |
| 2013, Koh TH [197] | Human | Surveillance |
| 2013, Lee GH [359] | Human | Therapeutics |
| 2013, Lee GH [456] | Human | Therapeutics |
| 2013, Lee LK [158] | Human | Public Health and Epidemiology |
| 2013, Li Y [96] | Human | Intervention |
| 2013, Liew YX [155] | Human | Public Health and Epidemiology |
| 2013, Lim CL-L [182] | Human | Public Health |
| 2013, Lim SG [376] | Human | Therapeutics |
| 2013, Ling ML [17] | Human, Environment | Transmission |
| 2013, Marimuthu K [457] | Human | Public Health and Epidemiology |
| 2013, Molton JS [188] | Human | Public Health and Epidemiology |
| 2013, Ng VWL [458] | Human | Therapeutics |
| 2013, Ong ZY [103] | Human | Intervention |
| 2013, Qiu G [123] | Environment | Intervention |
| 2013, Russell B [459] | Human | Diagnostics |
| 2013, Samy RP [460] | Human | Therapeutics |
| 2013, Samy RP [461] | Human | Therapeutics |
| 2013, Sarathy J [404] | Human | Microbiology |
| 2013, Sarathy J [403] | Human | Microbiology |
| 2013, Seah J [380] | Human | Therapeutics |

| Article ID | Sector(s) | Research Domain |
|---|---|---|
| 2013, Shao Q [73] | Human | Diagnostics |
| 2013, Sim JHC [159] | Human | Public Health and Epidemiology |
| 2013, Soon MML [12] | Human | Social and Economic Impacts |
| 2013, Tan PS [369] | Human | Therapeutics |
| 2013, Tan SY-Y [398] | Human | Microbiology |
| 2013, Tan TY [15] | Human, Environment | Transmission |
| 2013, Teo JQM [211] | Human | Surveillance |
| 2013, Teo JWP [198] | Human | Surveillance |
| 2013, Teo JWP [251] | Human | Surveillance |
| 2013, Vasudevan A [366] | Human | Therapeutics |
| 2013, Vasudevan A [156] | Human | Public Health and Epidemiology |
| 2013, Verrall A [179] | Human | Public Health and Epidemiology |
| 2013, Win M-K [55] | Human | Diagnostics |
| 2013, Xia E [255] | Human | Surveillance |
| 2013, Xiong P [462] | Human | Intervention |
| 2013, Yang Y [463] | Human | Therapeutics |
| 2013, Yokokawa F [317] | Human | Therapeutics |
| 2013, Yuan X [464] | Human | Intervention |
| 2013, Zou H [465] | Human | Therapeutics |
| 2014, Bandyopadhyay S [346] | Human | Therapeutics |
| 2014, Bayen S [75] | Environment | Diagnostics |
| 2014, Castillo CFG [180] | Human | Public Health and Epidemiology |
| 2014, Chen HH [466] | Human | Therapeutics |
| 2014, Ch'ng J-H [285] | Human | Therapeutics |
| 2014, Chow JY [467] | Human | Intervention |
| 2014, Coady DJ [468] | Human | Therapeutics |
| 2014, Deng CL [469] | Human | Therapeutics |
| 2014, Deng Y [470] | Human | Therapeutics |
| 2014, Gopal P [471] | Human | Therapeutics |
| 2014, Hon PY [133] | Human | Public Health and Epidemiology |
| 2014, Jauneikaite E [206] | Human | Surveillance |
| 2014, Khara JS [472] | Human | Therapeutics |
| 2014, Koh TH [207] | Human | Surveillance |
| 2014, La M-V [243] | Human | Surveillance |
| 2014, Lescar J [338] | Human | Therapeutics |
| 2014, Li X [102] | Human | Intervention |
| 2014, Liu SQ [473] | Human | Therapeutics |
| 2014, Loh CCY [474] | Human | Diagnostics |
| 2014, Mani V [475] | Human | Diagnostics |
| 2014, Marimuthu K [245] | Human | Surveillance |
| 2014, Marimuthu K [80] | Human | Intervention |
| 2014, Marimuthu K [87] | Human | Intervention |
| 2014, Mok S [341] | Human | Therapeutics |
| 2014, Ng TM [112] | Human | Intervention |
| 2014, Ng TM [163] | Human | Public Health and Epidemiology |
| 2014, Ng VWL [476] | Human | Therapeutics |
| 2014, Ong ZY [477] | Human | Therapeutics |
| 2014, Samy RP [478] | Human | Therapeutics |
| 2014, Seah XFV [108] | Human | Intervention |
| 2014, Shekar S [350] | Human | Therapeutics |
| 2014, Singhal A [313] | Human | Therapeutics |
| 2014, Song M [479] | Human | Therapeutics |
| 2014, Tan MW [181] | Human | Public Health and Epidemiology |
| 2014, Tan TT [16] | Human, Environment | Transmission |
| 2014, Tan YE [49] | Human | Diagnostics |
| 2014, Tang SS [186] | Human | Public Health and Epidemiology |
| 2014, Teo JWP [26] | Human, Environment | Transmission |
| 2014, Teo JWP [139] | Human | Public Health and Epidemiology |

| Article ID | Sector(s) | Research Domain |
|---|---|---|
| 2014, Vasudevan A [480] | Human | Diagnostics |
| 2014, Venkatachalam I [161] | Human | Public Health and Epidemiology |
| 2014, Wang Y [481] | Human | Therapeutics |
| 2014, Wen H [393] | Human | Microbiology |
| 2014, Wozniak M [482] | Human | Diagnostics |
| 2014, Wu Z [401] | Human | Microbiology |
| 2014, Yeoh LY [162] | Human | Public Health and Epidemiology |
| 2014, Young BE [170] | Human | Public Health and Epidemiology |
| 2014, Zhou YP [28] | Human | Transmission |
| 2014, Zhou YP [371] | Human | Therapeutics |
| 2015, Anusha S [284] | Human | Therapeutics |
| 2015, Cai Y [39] | Human | Diagnostics |
| 2015, Chee CBE [25] | Human | Transmission |
| 2015, Cheng J [286] | Human | Therapeutics |
| 2015, Cherng BPZ [191] | Human | Public Health and Epidemiology |
| 2015, Ch'ng JH [483] | Human | Microbiology |
| 2015, Choudhury S [365] | Human | Therapeutics |
| 2015, Chow ALP [3] | Human | Knowledge, Attitudes, Practices |
| 2015, Chow ALP [117] | Human | Intervention |
| 2015, Chua AP-G[362] | Human | Therapeutics |
| 2015, Chua NG [374] | Human | Therapeutics |
| 2015, Chung SJ [484] | Human | Therapeutics |
| 2015, Duan R [485] | Human | Therapeutics |
| 2015, Feng G [486] | Human | Therapeutics |
| 2015, Gopal P [326] | Human | Therapeutics |
| 2015, Harris PNA [358] | Human | Therapeutics |
| 2015, Harris PNA [487] | Human | Therapeutics |
| 2015, Harris PNA[90] | Human | Intervention |
| 2015, Haver HL [331] | Human | Therapeutics |
| 2015, Ho HJ [77] | Human | Intervention |
| 2015, Hsu L-Y [20] | Human | Transmission |
| 2015, Koh J-J [488] | Human | Therapeutics |
| 2015, Koh J-J [290] | Human | Therapeutics |
| 2015, Koh TH [236] | Human, Environment | Surveillance |
| 2015, Kumar A [384] | Human, Environment | Microbiology |
| 2015, Lau QY [310] | Human | Therapeutics |
| 2015, Lau QY [489] | Human | Therapeutics |
| 2015, Lau QY [490] | Human | Therapeutics |
| 2015, Lee HK [208] | Human | Surveillance |
| 2015, Lew KY [111] | Human | Intervention |
| 2015, Li J [491] | Human | Therapeutics |
| 2015, Liew YX [115] | Human | Intervention |
| 2015, Liew YX [11] | Human | Social and Economic Impacts |
| 2015, Lim K [104] | Human | Intervention |
| 2015, Lim T-P [348] | Human | Therapeutics |
| 2015, Lim T-P [492] | Human | Therapeutics |
| 2015, Ling ML [172] | Human | Public Health and Epidemiology |
| 2015, Loo LW [118] | Human | Intervention |
| 2015, Lu S [493] | Human | Intervention |
| 2015, Manjunatha UH [311] | Human | Therapeutics |
| 2015, Ng C [231] | Environment | Surveillance |
| 2015, Ng LSY [494] | Human | Public Health and Epidemiology |
| 2015, Ng PS [495] | Human | Therapeutics |
| 2015, Ng TM [375] | Human | Therapeutics |
| 2015, Paton NI [382] | Human | Therapeutics |
| 2015, Phoon YW [246] | Human | Surveillance |
| 2015, Samy RP [299] | Human | Therapeutics |
| 2015, Samy RP [496] | Human | Therapeutics |

| Article ID | Sector(s) | Research Domain |
|---|---|---|
| 2015, Seneviratne CJ [247] | Human | Surveillance |
| 2015, Tan S [300] | Human | Therapeutics |
| 2015, Tang SS [210] | Human | Surveillance |
| 2015, Teng CB [110] | Human | Intervention |
| 2015, Teo JWP [252] | Human | Surveillance |
| 2015, Vasoo S [190] | Human | Public Health and Epidemiology |
| 2015, Vasoo S [497] | Human | Therapeutics |
| 2015, Vasudevan A [7] | Human | Social and Economic Impacts |
| 2015, Win M-K [10] | Human | Social and Economic Impacts |
| 2015, Yi X [76] | Environment | Diagnostics |
| 2015, Yoon BK [345] | Human | Therapeutics |
| 2016, Ang TL [256] | Human | Surveillance |
| 2016, Ariyasu S [70] | Human | Diagnostics |
| 2016, Arora S [498] | Human | Therapeutics |
| 2016, Ashajyothi C [499] | Human | Therapeutics |
| 2016, Aung KT [220] | Food | Surveillance |
| 2016, Aung T-T [500] | Human | Therapeutics |
| 2016, Boudhar A [501] | Human | Therapeutics |
| 2016, Boudhar A [502] | Human | Therapeutics |
| 2016, Cai B [356] | Human | Therapeutics |
| 2016, Cai Y [107] | Human | Intervention |
| 2016, Cai Y [48] | Human | Diagnostics |
| 2016, Cai Y [305] | Human | Therapeutics |
| 2016, Cao Y [503] | Human | Therapeutics |
| 2016, Chen YT [364] | Human | Therapeutics |
| 2016, Chen Y-T [31] | Human | Transmission |
| 2016, Chia G [84] | Human | Intervention |
| 2016, Chow ALP [143] | Human | Public Health and Epidemiology |
| 2016, Chow ALP [116] | Human | Intervention |
| 2016, Chua SL [504] | Human | Therapeutics |
| 2016, Fisher D [93] | Human, Environment | Intervention |
| 2016, Ghode P [325] | Human | Therapeutics |
| 2016, Gopal P [329] | Human | Therapeutics |
| 2016, Hemu X [505] | Human | Therapeutics |
| 2016, Ho HJ [21] | Human | Transmission |
| 2016, Husain N [506] | Human | Therapeutics |
| 2016, Isenman H [85] | Human | Intervention |
| 2016, Isenman H [86] | Human | Intervention |
| 2016, Khara JS [507] | Human | Therapeutics |
| 2016, Khong WX [29] | Human | Transmission |
| 2016, Khong WX [32] | Human | Transmission |
| 2016, Koh J-J [508] | Human | Therapeutics |
| 2016, Kong YL [509] | Human | Public Health and Epidemiology |
| 2016, Lakshminarayanan R [336] | Human | Therapeutics |
| 2016, Le T-H [227] | Environment | Surveillance |
| 2016, Lee SH [510] | Human | Therapeutics |
| 2016, Li M [511] | Human | Therapeutics |
| 2016, Lim EJZ [223] | Food | Surveillance |
| 2016, Lim MY-X [339] | Human | Therapeutics |
| 2016, Lim WS [512] | Human | Therapeutics |
| 2016, Low A [228] | Environment | Surveillance |
| 2016, Mandakhalikar KD [98] | Human | Intervention |
| 2016, Marimuthu K [94] | Human | Intervention |
| 2016, Ng TM [361] | Human | Therapeutics |
| 2016, Pan DST [4] | Human | Knowledge, Attitudes, Practices |
| 2016, Seneviratne CJ [396] | Human | Microbiology |
| 2016, Song CT [153] | Human | Public Health and Epidemiology |
| 2016, Su CT-T [352] | Human | Therapeutics |

| **Article ID** | **Sector(s)** | **Research Domain** |
|---|---|---|
| 2016, Teng CP [513] | Human | Therapeutics |
| 2016, Teo JQM [185] | Human | Public Health and Epidemiology |
| 2016, Teo JWP [199] | Human | Surveillance |
| 2016, Teo JWP [53] | Human | Diagnostics |
| 2016, Tran NH [74] | Environment | Diagnostics |
| 2016, Tran NH [121] | Environment | Intervention |
| 2016, Truong T [514] | Human | Therapeutics |
| 2016, Tun ZM [5] | Human | Knowledge, Attitudes, Practices |
| 2016, Wee KB [354] | Human | Therapeutics |
| 2016, Wong EHH [515] | Human | Therapeutics |
| 2016, Wong JG [178] | Human | Public Health and Epidemiology |
| 2016,Teo JQM [249] | Human | Surveillance |
| 2017 Lee MHM [516] | Human | Intervention |
| 2017 Zhang J [517] | Food, Environment | Intervention |
| 2017, Ang MLT [318] | Human | Therapeutics |
| 2017, Aung KT [235] | Food, Environment | Surveillance |
| 2017, Aw J [71] | Human | Diagnostics |
| 2017, Aziz DB [377] | Human | Therapeutics |
| 2017, Cai Y [214] | Human | Surveillance |
| 2017, Cai Y [304] | Human | Therapeutics |
| 2017, Chee CBE [378] | Human | Therapeutics |
| 2017, Chen S [518] | Human | Diagnostics |
| 2017, Chew KL [144] | Human | Public Health and Epidemiology |
| 2017, Chew KL [129] | Human | Public Health and Epidemiology |
| 2017, Chew KL [184] | Human | Public Health |
| 2017, Chew KL [43] | Human | Diagnostics |
| 2017, Chin JSF [519] | Human | Diagnostics |
| 2017, Chong SM [221] | Food, Animal | Surveillance |
| 2017, Chow ALP [19] | Human | Transmission |
| 2017, Gopal P [327] | Human | Therapeutics |
| 2017, Gopal P [328] | Human | Therapeutics |
| 2017, Hou Z [288] | Human | Therapeutics |
| 2017, Hsu L-Y [130] | Human | Public Health and Epidemiology |
| 2017, Hsu L-Y [27] | Human, Animal | Transmission |
| 2017, Khara JS [520] | Human | Therapeutics |
| 2017, Kityo Cissy [521] | Human | Therapeutics |
| 2017, Lee T-H [1] | Human | Knowledge, Attitudes, Practices |
| 2017, Li PQ [522] | Human | Diagnostics |
| 2017, Lin S [291] | Human | Therapeutics |
| 2017, Lin S [292] | Human | Therapeutics |
| 2017, Lin S [293] | Human | Therapeutics |
| 2017, Liu S [294] | Human | Therapeutics |
| 2017, Loo LH [146] | Human | Public Health and Epidemiology |
| 2017, Marimuthu K [131] | Human | Public Health and Epidemiology |
| 2017, Ng C [232] | Environment | Surveillance |
| 2017, Ng SMS [523] | Human | Therapeutics |
| 2017, Ng SMS [524] | Human | Therapeutics |
| 2017, Ng SMS [525] | Human | Therapeutics |
| 2017, Paton NI [68] | Human | Diagnostics |
| 2017, Peng J [349] | Human | Therapeutics |
| 2017, Pu Y [526] | Human | Therapeutics |
| 2017, Rashid R [399] | Human | Microbiology |
| 2017, Samy RP [527] | Human | Therapeutics |
| 2017, Seneviratne CJ [528] | Human | Therapeutics |
| 2017, Tan JH [529] | Human | Therapeutics |
| 2017, Tan JPK [530] | Human | Therapeutics |
| 2017, Tan TY [160] | Human | Public Health and Epidemiology |
| 2017, Tan TY [57] | Human | Diagnostics |

| Article ID | Sector(s) | Research Domain |
|---|---|---|
| 2017, Teo JQM [164] | Human | Public Health and Epidemiology |
| 2017, Venkatesh M [301] | Human | Therapeutics |
| 2017, Wang A [531] | Human | Microbiology |
| 2017, Wang J [532] | Human | Therapeutics |
| 2017, Wang K [395] | Human | Microbiology |
| 2017, Yee M [344] | Human | Therapeutics |
| 2017, Yew PYM [533] | Human | Therapeutics |
| 2017, Yi X [119] | Environment | Intervention |
| 2017, Zhong G [303] | Human | Therapeutics |
| 2017, Zhou C [101] | Human | Intervention |
| 2018, Aziz DB [308] | Human | Therapeutics |
| 2018, Baek JS [534] | Human, Environment | Therapeutics |
| 2018, Balne PK [99] | Human | Intervention |
| 2018, Budigi Y [535] | Human | Therapeutics |
| 2018, Cai Q [536] | Environment | Intervention |
| 2018, Cai Y [38] | Human | Diagnostics |
| 2018, Chan HL [537] | Human | Diagnostics |
| 2018, Chan LY [69] | Human | Diagnostics |
| 2018, Chan MKL [51] | Human | Diagnostics |
| 2018, Chew KL [52] | Human | Diagnostics |
| 2018, Chew KL [355] | Human | Therapeutics |
| 2018, Chiang RZ-H [347] | Human | Therapeutics |
| 2018, Chiew CJ [167] | Human | Public Health and Epidemiology |
| 2018, Chilambi GS [538] | Human | Therapeutics |
| 2018, Chin W [322] | Human | Therapeutics |
| 2018, Chow ALP [88] | Human | Intervention |
| 2018, Chuang L [539] | Human | Therapeutics |
| 2018, Ding Y [33] | Human | Transmission |
| 2018, Ding Y [385] | Food | Microbiology |
| 2018, Gao J [540] | Human | Diagnostics |
| 2018, Haller L [226] | Environment | Surveillance |
| 2018, Hartantyo SHP [541] | Animal | Surveillance |
| 2018, Heng ST [542] | Human | Public Health and Epidemiology |
| 2018, Ho ZJM [23] | Human | Transmission |
| 2018, Htun JL [149] | Human | Public Health and Epidemiology |
| 2018, Hu B [543] | Human | Therapeutics |
| 2018, Kalimuddin S [383] | Human | Therapeutics |
| 2018, Koh JJ [544] | Human | Therapeutics |
| 2018, Koh TH [391] | Human | Microbiology |
| 2018, Lakshimarayanan R [545] | Human | Therapeutics |
| 2018, Lau QY [337] | Human | Therapeutics |
| 2018, Le T-H [122] | Environment | Intervention |
| 2018, Lee G-H [64] | Human | Diagnostics |
| 2018, Li J [546] | Human | Therapeutics |
| 2018, Lim CLL [168] | Human | Public Health and Epidemiology |
| 2018, Lim T-P [381] | Human | Therapeutics |
| 2018, Lim YH [547] | Human | Therapeutics |
| 2018, Lou W [295] | Human | Therapeutics |
| 2018, Mendez AR [97] | Human, Environment | Intervention |
| 2018, Ng C [229] | Environment | Surveillance |
| 2018, Ng C [230] | Environment | Surveillance |
| 2018, Ng C [388] | Environment | Microbiology |
| 2018, Ng DHL [14] | Human, Environment | Transmission |
| 2018, Ng SMS [312] | Human | Therapeutics |
| 2018, Ng SMS [548] | Human | Therapeutics |
| 2018, Obuobi S [549] | Human | Therapeutics |
| 2018, Ong CH [41] | Human | Diagnostics |
| 2018, Parmar A [550] | Human | Therapeutics |

| Article ID | Sector(s) | Research Domain |
|---|---|---|
| 2018, Rocamora F [402] | Human | Microbiology |
| 2018, Selcuk A [217] | Human | Surveillance |
| 2018, Sinha S [551] | Human | Therapeutics |
| 2018, Su CT-T [351] | Human | Therapeutics |
| 2018, Su W [406] | Human | Microbiology |
| 2018, Tan BH [363] | Human | Therapeutics |
| 2018, Tan D [169] | Human | Public Health and Epidemiology |
| 2018, Tan YE [248] | Human | Surveillance |
| 2018, Tang YW [552] | Human | Therapeutics |
| 2018, Teo JWP [390] | Human | Microbiology |
| 2018, Teo JWP [58] | Human | Diagnostics |
| 2018, Tong JX [315] | Human | Therapeutics |
| 2018, Xu HV [553] | Human | Intervention |
| 2018, Yang SS [554] | Human | Therapeutics |
| 2018, Yason JA [316] | Human | Therapeutics |
| 2018, Yeo CK [555] | Human | Therapeutics |
| 2018, Yuan W [126] | Food | Intervention |
| 2018, Zhang Z [72] | Human | Diagnostics |
| 2018, Zhu L [556] | Human | Public Health and Epidemiology |
| 2018, Zwe YH [225] | Food | Surveillance |
| 2019, Arfan G [557] | Human | Therapeutics |
| 2019, Aung KT [219] | Food, Animal | Surveillance |
| 2019, Bae K [558] | Human | Diagnostics |
| 2019, Bazan EL [559] | Human | Therapeutics |
| 2019, Bharadwaj S [91] | Human | Intervention |
| 2019, Chan JC [367] | Human | Therapeutics |
| 2019, Chan YY [2] | Human | Knowledge, Attitudes, Practices |
| 2019, Chen H [34] | Human, Environment | Transmission |
| 2019, Chen WK [560] | Human | Public Health and Epidemiology |
| 2019, Chew KL [30] | Human | Transmission |
| 2019, Chew KL [67] | Human | Diagnostics |
| 2019, Chiu JKH [561] | Human | Diagnostics |
| 2019, Choudhury S [47] | Human | Diagnostics |
| 2019, Chua AQ [193] | Human | Public Health and Epidemiology |
| 2019, Dupont C [324] | Human | Therapeutics |
| 2019, Ero R [562] | Human | Therapeutics |
| 2019, Fong J [563] | Human | Intervention |
| 2019, Guo S [222] | Food | Surveillance |
| 2019, Guo S [386] | Food | Microbiology |
| 2019, Guo S [389] | Food | Microbiology |
| 2019, Ho HJ [154] | Human | Public Health and Epidemiology |
| 2019, Ho PL [332] | Human | Therapeutics |
| 2019, Htun HL [148] | Human | Public Health and Epidemiology |
| 2019, Keerthisinghe TP [564] | Environment | Intervention |
| 2019, Ko KKK [240] | Human | Surveillance |
| 2019, Kyaw BM [565] | Human | Intervention |
| 2019, La M-V [138] | Human | Public Health and Epidemiology |
| 2019, Lee SY [42] | Human | Diagnostics |
| 2019, Legido-Quigley H [194] | Human | Public Health and Epidemiology |
| 2019, Leung CM [95] | Human | Intervention |
| 2019, Li D [127] | Food | Intervention |
| 2019, Li M [566] | Human | Therapeutics |
| 2019, Liu H [392] | Environment | Microbiology |
| 2019, Loke HY [176] | Human | Public Health and Epidemiology |
| 2019, Loo LW [106] | Human | Intervention |
| 2019, Marimuthu K [173] | Human | Public Health and Epidemiology |
| 2019, Martinez-Vega R [567] | Human | Public Health and Epidemiology |
| 2019, Mayandi V [296] | Human | Therapeutics |

| Article ID | Sector(s) | Research Domain |
|---|---|---|
| 2019, Mo Y [6] | Human | Knowledge, Attitudes, Practices |
| 2019, Mo Y [13] | Human | Social and Economic Impacts |
| 2019, Ng C [120] | Environment | Intervention |
| 2019, Ngo T-M [568] | Human | Diagnostics |
| 2019, Obuobi, S [569] | Human | Therapeutics |
| 2019, Octavia S [136] | Human | Public Health and Epidemiology |
| 2019, Pada, SMSK [81] | Human | Intervention |
| 2019, Pang X [128] | Food | Intervention |
| 2019, Quek WM [372] | Human | Therapeutics |
| 2019, Ryanputra D [373] | Human | Therapeutics |
| 2019, Selcuk A [218] | Human | Surveillance |
| 2019, Singh SR [195] | Human | Public Health and Epidemiology |
| 2019, Sorayah R [342] | Human | Therapeutics |
| 2019, Stewardson AJ [166] | Human | Public Health and Epidemiology |
| 2019, Su CT-T [570] | Human | Therapeutics |
| 2019, Tan GSE [36] | Human | Transmission |
| 2019, Tan YE [135] | Human | Public Health and Epidemiology |
| 2019, Tay MYF [224] | Food, Human | Surveillance |
| 2019, Teo JQM [343] | Human | Therapeutics |
| 2019, Teo JQM [571] | Human | Public Health and Epidemiology |
| 2019, Teo SW [572] | Human | Therapeutics |
| 2019, Tong JX [314] | Human | Therapeutics |
| 2019, Tran NH [573] | Environment | Surveillance |
| 2019, Vasoo S [59] | Human | Diagnostics |
| 2019, Yang C [302] | Human | Therapeutics |
| 2019, Yi X [233] | Environment | Surveillance |
| 2019, Yi X [234] | Environment | Surveillance |
| 2019, Yuan Y [574] | Human | Therapeutics |
| 2019, Yuan Y [575] | Human | Therapeutics |
| 2019, Zhang EX [183] | Human | Public Health and Epidemiology |
| 2019, Zhang J [576] | Food, Environment | Intervention |
| 2019, Zhang K [577] | Human | Therapeutics |
| 2019, Zhang L [578] | Environment | Intervention |
| 2019, Zheng S-W [579] | Human | Public Health and Epidemiology |
| 2019, Zhou C [100] | Human | Intervention |

**Supplementary Table ST2. List of AMR research or review articles published between 2009 and 2019 where Singapore either contributed samples or data, or were involved as collaborators.**

Articles were assigned to the most relevant research domain, sector(s) and the lead or collaborating country.

| Article ID | Lead or collaborating country | Sector(s) | Research Domain |
|---|---|---|---|
| **Contributing samples to multinational studies (SGS), 2009-2019** | | | |
| 2009, Bouchillon SK [258] | USA | Human | Surveillance |
| 2009, Chuang C-H [580] | Taiwan | Human | Surveillance |
| 2009, Hawser SP [264] | Switzerland | Human | Surveillance |
| 2009, Hurt AC [266] | Australia | Human | Surveillance |
| 2009, Ko W-C [581] | Taiwan | Human | Surveillance |
| 2009, Lee H-Y [273] | Taiwan<br>South Korea | Human | Surveillance |
| 2009, Mendes RE [582] | USA | Human | Surveillance |
| 2009, Yau W [283] | Australia | Human | Surveillance |
| 2010, Christiansen KJ [261] | UK | Human | Surveillance |
| 2010, Farrell DJ [262] | USA | Human | Surveillance |
| 2010, Higgins PG [583] | Germany | Human | Public Health and Epidemiology |
| 2010, Hsueh P-R [265] | Taiwan | Human | Surveillance |
| 2011, Chen Y-H [260] | Taiwan | Human | Surveillance |
| 2011, Chung DR [584] | South Korea | Human | Public Health and Epidemiology |
| 2011, Hurt AC [267] | Australia | Human | Surveillance |
| 2011, Hurt AC [585] | Australia | Human | Public Health and Epidemiology |
| 2011, Lee MY [586] | South Korea | Human | Public Health and Epidemiology |
| 2011, Roberts JA [587] | USA | Human | Therapeutics |
| 2011, Wang H [282] | China<br>Taiwan | Human | Surveillance |
| 2012, Bouchillon S [257] | USA | Human | Surveillance |
| 2012, Kiratisin P [271] | Thailand | Human | Surveillance |
| 2012, Lin Y-T [588] | Taiwan | Human | Public Health and Epidemiology |
| 2012, Lu P-L [274] | Taiwan | Human | Surveillance |
| 2012, Namdari H [276] | USA | Human | Surveillance |
| 2012, Yang Y [589] | China (Hong Kong SAR) | Environment | Surveillance |
| 2013, Holden MTG [590] | UK<br>Germany<br>Ireland | Human | Public Health and Epidemiology |
| 2013, Kim DH [591] | South Korea | Human | Transmission |
| 2013, Leang S-K [272] | Australia | Human | Surveillance |
| 2013, Mendes RE [275] | USA | Human | Surveillance |
| 2013, Sader HS [279] | USA | Human | Surveillance |
| 2013, Sheng W-H [592] | Taiwan | Human | Surveillance |
| 2014, Ginn AN [593] | Australia | Human | Diagnostics |
| 2015, Holt KE [594] | Australia<br>UK | Human,<br>Animal | Public Health and Epidemiology |
| 2015, Pfaller MA [277] | USA | Human | Surveillance |
| 2016, Jean S-S [268] | Taiwan | Human | Surveillance |
| 2016, Tan TY [280] | Singapore | Human | Surveillance |
| 2016, Torumkuney D [281] | UK | Human | Surveillance |
| 2017, Blackwell GA [595] | Australia | Human | Transmission |
| 2017, Blackwell GA [596] | Australia | Human | Microbiology |
| 2017, Chang Y-T [259] | Taiwan | Human | Surveillance |
| 2017, Cheong HS [597] | South Korea | Human | Surveillance |
| 2017, Jean S-S [269] | Taiwan | Human | Surveillance |
| 2017, Karlowsky JA [270] | US | Human | Surveillance |
| 2017, Ma L [598] | China (Hong Kong SAR) | Environment | Surveillance |
| 2018, Harris PNA [599] | Australia | Human | Public Health and Epidemiology |
| 2018, Harris PNA [600] | Australia | Human | Therapeutics |
| 2018, Khor WC [601] | Malaysia | Human | Public Health and Epidemiology |
| 2018, Mendis SM [602] | Singapore<br>USA | Human | Public Health and Epidemiology |

| Article ID | Lead or collaborating country | Sector(s) | Research Domain |
|---|---|---|---|
| 2018, Pfaller MA [278] | USA | Human | Surveillance |
| 2018, Versporten A [603] | Belgium | Human | Surveillance |
| 2018, Vilaichone RK [604] | Thailand | Human | Public Health and Epidemiology |
| 2018, Yarbrough ML [605] | USA | Human | Diagnostics |
| 2019, Blot S [606] | Belgium | Human | Public Health and Epidemiology |
| 2019, Chen SL [607] | Singapore | Human | Public Health and Epidemiology |
| 2019, George CRR [263] | Australia | Human | Surveillance |
| 2019, Hendriksen RS [608] | Denmark | Environment | Surveillance |
| 2019, Hsia Y [609] | UK | Human | Surveillance |
| 2019, Hsia Y [610] | UK | Human | Surveillance |
| 2019, Hu YJ [611] | China (Hong Kong SAR) | Human | Public Health and Epidemiology |
| 2019, Luo Y [612] | China<br>USA | Human | Public Health and Epidemiology |
| 2019, Ma L [613] | China (Hong Kong SAR) | Environment | Surveillance |
| 2019, Papadopoulos A [614] | Greece | Human | Public Health and Epidemiology |
| **International Research Collaborations on AMR (SGO), 2009-2019** | | | |
| 2009, Cervantes S [615] | USA | Human | Diagnostics |
| 2009, Tam VH [616] | USA | Human | Therapeutics |
| 2009, Valvatne H [617] | Norway | Human | Public Health and Epidemiology |
| 2011, Dhorda M [618] | France | Human | Transmission |
| 2011, Leitsch D [619] | Australia | Human | Therapeutics |
| 2011, Massi MN [620] | Indonesia | Human | Public Health and Epidemiology |
| 2011, Siswantoro H [621] | Australia | Human | Therapeutics |
| 2012, Apisarnthanarak A [622] | Thailand | Human, Environment | Public Health and Epidemiology |
| 2012, Brunner R [623] | Switzerland | Human | Therapeutics |
| 2012, Cervantes S [624] | USA | Human | Diagnostics |
| 2012, Dunn LA [625] | Australia | Human | Therapeutics |
| 2012, Guiton PS [626] | USA | Human | Intervention |
| 2012, Ho KKK [627] | Australia | Human | Intervention |
| 2012, Köser CU [628] | UK | Human | Diagnostics |
| 2012, Lee M [629] | South Korea | Human | Therapeutics |
| 2012, Shields RK [630] | USA | Human | Public Health and Epidemiology |
| 2012, Zhang L [631] | China | Human | Microbiology |
| 2013, Barraud N [632] | Australia | Human | Therapeutics |
| 2013, Bowers DR [633] | Singapore<br>USA | Human | Therapeutics |
| 2013, Chiang W-C [634] | Denmark | Human | Therapeutics |
| 2013, Ciesielczuk H [635] | UK | Human | Diagnostics |
| 2013, Jakobsen TH [636] | Denmark | Human | Microbiology |
| 2013, Kelesidis T [637] | USA | Human | Surveillance |
| 2013, Liu Y [638] | Denmark | Human | Therapeutics |
| 2013, Richmond GE [639] | UK | Human | Diagnostics |
| 2013, Wang H [640] | Denmark | Human | Therapeutics |
| 2013, Yepuri NR [641] | Australia | Human | Intervention |
| 2013, Zhang Y [642] | USA | Animal, Environment | Surveillance |
| 2013, Zhang Y [643] | USA | Animal, Environment | Surveillance |
| 2014, Apisarnthanarak A [644] | Thailand | Human | Intervention |
| 2014, Butler J [645] | Australia | Human | Public Health and Epidemiology |
| 2014, Duan S [646] | USA | Human | Microbiology |
| 2014, Karunakaran R [647] | Malaysia | Human | Public Health and Epidemiology |
| 2014, Kelesidis T [648] | USA | Human, Animal | Transmission |
| 2014, Lai C-C [649] | Taiwan | Human | Public Health and Epidemiology |
| 2014, Landelle C [650] | Switzerland | Human, Environment | Intervention |

| Article ID | Lead or collaborating country | Sector(s) | Research Domain |
|---|---|---|---|
| 2014, Stryjewski ME [651] | Argentina | Human | Therapeutics |
| 2014, Veiga MI [652] | Portugal | Human | Microbiology |
| 2015, Aamodt H [653] | Norway | Human | Public Health and Epidemiology |
| 2015, Baranovich T [654] | USA | Human | Therapeutics |
| 2015, Farrukee R [655] | Australia | Human | Therapeutics |
| 2015, Malmquist NA [656] | France | Human | Therapeutics |
| 2015, Matsunaga S [657] | Japan | Human | Diagnostics |
| 2015, Nguyen D [658] | Australia | Human | Therapeutics |
| 2015, Regmi SM [659] | Thailand | Human | Surveillance |
| 2015, Rhee S-Y [660] | USA | Human | Diagnostics |
| 2015, Soetaert K [661] | Belgium | Human | Therapeutics |
| 2015, Zowawi HM [662] | Australia | Human | Public Health and Epidemiology |
| 2016, Arribas JR [663] | Spain | Human | Therapeutics |
| 2016, Auburn S [664] | Australia | Human | Diagnostics |
| 2016, Coker OO [665] | Thailand | Human | Public Health and Epidemiology |
| 2016, Cunningham SA [666] | USA | Human | Diagnostics |
| 2016, Grigg MJ [667] | Australia | Human | Therapeutics |
| 2016, Hanafi A [668] | Malaysia | Human | Microbiology |
| 2016, Harris RC [669] | UK | Human | Therapeutics |
| 2016, Mac Aogáin M [670] | Ireland | Human | Surveillance |
| 2016, Nguyen TK [671] | Australia | Human | Therapeutics |
| 2016, Nilsson M [672] | Denmark | Human | Microbiology |
| 2016, Phyo AP [673] | Thailand | Human | Therapeutics |
| 2016, Richmond GE [674] | UK | Human | Microbiology |
| 2016, Stenvang M [675] | Denmark | Human | Therapeutics |
| 2017, Atarashi K [676] | Japan | Human | Microbiology |
| 2017, Basilico N [677] | Italy | Human | Therapeutics |
| 2017, Bazaka K [678] | Australia | Human | Therapeutics |
| 2017, Belousoff MJ [679] | Australia | Human | Therapeutics |
| 2017, Bhuyan GS [680] | Bangladesh | Human | Public Health and Epidemiology |
| 2017, Chang MJ [681] | Republic of Korea (South Korea) | Human | Therapeutics |
| 2017, Chen F [682] | China | Human | Therapeutics |
| 2017, Cunningham SA [683] | USA | Human | Diagnostics |
| 2017, Gowrisankar G [684] | India | Environment | Microbiology |
| 2017, Gunawan C [685] | Australia | Human | Intervention |
| 2017, Howlin RP [686] | UK | Human | Therapeutics |
| 2017, Hutchison C [687] | China | Human | Social and Economic Impacts |
| 2017, Kathirvel S [688] | India | Human | Knowledge, Attitudes, Practices |
| 2017, Lamoth F [689] | USA | Human | Public Health and Epidemiology |
| 2017, Landier J [690] | Thailand | Human | Therapeutics |
| 2017, Liao J-H [691] | Taiwan | Human | Microbiology |
| 2017, Thai VC [692] | Vietnam | Human | Therapeutics |
| 2018, Ahmed W [693] | Australia | Environment | Surveillance |
| 2018, Antonoplis A [694] | USA | Human | Therapeutics |
| 2018, Beattie RE [695] | USA | Environment | Surveillance |
| 2018, Chen C [696] | USA | Human | Therapeutics |
| 2018, CRyPTIC Consortium [697] | UK | Human | Diagnostics |
| 2018, Dunn DT [698] | UK | Human | Diagnostics |
| 2018, Fang T [699] | China | Environment | Surveillance |
| 2018, Germond A [700] | Japan | Human | Microbiology |
| 2018, Grigg MJ [701] | Australia | Human | Therapeutics |
| 2018, Hoppe A [702] | UK | Human | Therapeutics |
| 2018, Jiang Y [703] | China | Environment | Surveillance |
| 2018, Kano R [704] | Japan | Animal | Therapeutics |
| 2018, Malkawi R [705] | UK | Human | Therapeutics |
| 2018, Mather AE [706] | Vietnam | Human | Transmission |

| Article ID | Lead or collaborating country | Sector(s) | Research Domain |
|---|---|---|---|
| 2018, Merchant S [707] | USA | Human | Public Health and Epidemiology |
| 2018, Nordström R [708] | Sweden | Human | Therapeutics |
| 2018, Oonsivilai M [709] | Cambodia | Human | Diagnostics |
| 2018, Ravensdale JT [710] | Australia | Human | Surveillance |
| 2018, Stockdale AJ [711] | UK | Human | Therapeutics |
| 2018, Subedi D [712] | Australia | Human | Microbiology |
| 2018, Subedi D [713] | Australia | Human | Public Health and Epidemiology |
| 2018, Subedi D [714] | Australia | Human | Transmission |
| 2018, Tzou PL [715] | USA | Human | Diagnostics |
| 2018, Vente A [716] | Germany | Human | Therapeutics |
| 2018, Zhou C [717] | China | Human | Intervention |
| 2019, Brunton LA [718] | UK | Environment | Public Health and Epidemiology |
| 2019, Capci A [719] | Germany | Human | Therapeutics |
| 2019, Chen Y [720] | China | Environment | Surveillance |
| 2019, Chen Y [721] | China | Environment | Surveillance |
| 2019, Faksri K [722] | Thailand | Human | Diagnostics |
| 2019, González A [723] | Spain | Human | Therapeutics |
| 2019, González A [724] | Spain | Human | Therapeutics |
| 2019, Jabbar A [725] | Pakistan | Human | Public Health and Epidemiology |
| 2019, Juhas M [726] | Switzerland | Human | Therapeutics |
| 2019, Li H [727] | Denmark | Food | Transmission |
| 2019, Li H [728] | Denmark | Food | Surveillance |
| 2019, Limmathurotsakul D [729] | Thailand | Human, Animal, Food | Knowledge, Attitudes, Practices |
| 2019, Long S [730] | China | Human | Diagnostics |
| 2019, Nilsson M [731] | Denmark | Human | Therapeutics |
| 2019, Nilsson M [732] | Denmark | Human | Therapeutics |
| 2019, Pei M [733] | China | Environment | Intervention |
| 2019, Penesyan A [734] | Australia | Human | Therapeutics |
| 2019, Phelan JE [735] | UK<br>Philippines | Human | Microbiology |
| 2019, Ram M R [736] | Malaysia | Human | Therapeutics |
| 2019, Safi H [737] | USA | Human | Microbiology |
| 2019, Sosibo SC [738] | South Africa | Human | Therapeutics |
| 2019, Subedi D [739] | Australia | Human | Microbiology |
| 2019, Thompson JA [740] | UK | Human | Therapeutics |
| 2019, Yang DL [741] | China | Human | Therapeutics |
| 2019, Zhang N [742] | China | Environment | Surveillance |

**Supplementary Table ST3. Number of articles published by each group of institutions, 2009-2019.**

| Singapore Institutions (by groups) | Number of articles |
|---|---|
| Institutes of Higher Learning | 245 |
| Ministry / Agency | 13 |
| Research Institutes | 102 |
| Public Healthcare Institutions | 260 |

**Supplementary Table ST4. Summary of the types of AMR research that Singapore contributed samples or data to, 2009-2019**

| Domain | Sector | Types of studies | Endnote |
|---|---|---|---|
| Surveillance | Human | Most studies reported the resistance trends of microorganisms to antimicrobials. Some of these studies reported surveillance data from regional studies such as the Study for Monitoring Antimicrobial Resistance Trends (SMART), the COMPACT study, the Survey of Antibiotic Resistance (SOAR), the SENTRY antifungal surveillance programme, the Tigecycline Evaluation and Surveillance Trial (TEST), and the Community-Acquired Respiratory Tract Infection Pathogen Surveillance (CARTIPS) study. | [257-263, 265, 268, 270, 271, 273-283, 580, 581] |
| | | A few studies reported the trends in antimicrobial consumption in adults and children. | [603, 609, 610] |
| | | Emergence and distribution of ESBL, AmpC beta-lactamases and carbapenemases were reported. | [264, 269, 592, 597] |
| | | The emergence and spread of carbapenemase genes was reported in one study from the SENTRY Surveillance Programme. | [582] |
| | | Studies on the emergence and spread of influenza A(H1N1) towards antivirals. All studies identified focused after the 2009 pandemic when there was increased use of antivirals. | [266, 267, 272] |
| | Environment | Drinking water samples from households or point of use were collected to investigate the antibiotic resistome. | [598, 613] |
| | | AMR gene abundance from urban sewage or sewage treatment plants were studied to identify variations and diversity. | [589, 608] |
| Public Health and Epidemiology | Human | Studies in the human sector were diverse and consisted of investigations into the genetic linkages with resistance phenotypes. | [586, 599] |
| | | Elucidating resistance profile with pathogen characteristics. | [602, 607] |
| | | Correlating resistance profile with the type of antimicrobial treatment received. | [584, 614] |
| | | Population characteristics for the acquisition or colonisation by resistant microorganisms. | [588, 601, 606, 611, 612] |
| | | The pattern and spread of resistant *Helicobacter pylori*, carbapenem-resistance *Acinetobacter baumannii* and MRSA were studied. | [583, 590, 604] |
| | | One study on resistant influenza H1N1 isolated form patients receiving antiviral therapy. | [585] |
| | Human-animal | *Klebsiella pneumoniae* from human and animal sources were analysed to determine its diversity and population structure, as well as their virulence and AMR. | [594] |
| Diagnostics | Human | One study that identified gene targets that could be used to predict resistance to 3GC and aminoglycosides in *Klebsiella pneumoniae* and *Escherichia coli*.<br>Another study evaluated the performance of the Xpert MRSA NxG assay in detecting MRSA directly from nasal swabs. | [593, 605] |
| Therapeutics | Human | Clinical trial to investigate the activity of piperacillin-tazobactam and meropenem in patients with bacteraemia due to ceftriaxone-resistant *E. coli* or *K. pneumoniae*.<br>Pharmacodynamic simulation of carbapenem infusions to determine the dosing regimens required to achieve effective and optimal cumulative fraction of response against resistant bacteria. | [587, 600] |
| Transmission | Human | Genetic determinants of spread were investigated in *Acinetobacter baumannii*. | [591, 595] |
| Microbiology | Human | Presence of an *armA* gene (a 16S methyltransferase shown to confer resistance to several aminoglycosides) in carbapenem-resistant *Acinetobacter baumannii* was analysed. | [596] |

T, Fjeldsoee-Nielsen H, Kott M, Kostoula A, Javeri Y, Einav S, Makikado LDU, Tomescu D, Gritsan A, Jovanovic B, Venkatesan K, Mirkovic T, Emmerich M, Canale M, Dietz LS, Ilutovich S, Miñope JTS, Silva RB, Montenegro MA, Martin P, Saul P, Chediack V, Sutton G, Couce R, Balasini C, Gonzalez S, Lascar FM, Descotte EJ, Gumiela NS, Pino CA, Cesio C, Valgolio E, Cunto E, Dominguez C, Nelson NF, Abegao EM, Pozo NC, Bianchi L, Correger E, Pastorino ML, Miyazaki EA, Grubissich N, Garcia M, Bonetto N, Quevedo NE, Gomez CD, Queti F, Estevarena LG, Cruz G, Fernandez R, Santolaya I, Grangeat SH, Doglia J, Zakalik G, Pellegrini C, Lloria MM, Chacon ME, Fumale M, Leguizamon M, Hidalgo IB, Tiranti RJ, Capponi P, Tita A, Cardonnet L, Bettini L, Ramos A, Lovesio L, Miranda EM, Farfan AB, Tolosa C, Segura L, Bellocchio A, Alvarez B, Manzur A, Lujan R, Fernandez N, Scarone N, Zazu A, Groh C, Fletcher J, Smith J, Azad R, Chavan N, Wong H, Kol M, Campbell L, Starr T, Roberts B, Wibrow B, Warhurst T, Chinthamuneedi M, Ferney BB, Simon M, De Backer D, Wittebole X, De Bels D, Collin V, Dams K, Jorens P, Dubois J, Gunst J, Haentjens L, De Schryver N, Dugernier T, Rizoli S, Santillan P, Han Y, Biskup E, Qu C, Li X, Yu T, Lu W, Molano-Franco D, Rojas J, Oviedo JMP, Pinilla D, Celis E, Arias M, Vukovic A, Vudrag M, Belavic M, Zunic J, Kuharic J, Kricka IB, Filipovic-Grcic I, Tomasevic B, Obraz M, Bodulica B, Dohnal M, Malaska J, Kratochvil M, Satinsky I, Schwarz P, Kos Z, Blahut L, Maca J, Protus M, Kieslichova E, Nielsen LG, Krogh BM, Rivadeneira F, Morales F, Mora J, Orozco AS, MorochoTutillo DR, Vargas NR, Yepez ES, Villamagua B, Alsisi A, Fahmy A, Dupont H, Lasocki S, Paugam-Burtz C, Foucrier A, Nica A, Barjon G, Mallat J, Marcotte G, Leone M, Duclos G, Burtin P, Atchade E, Mahjoub Y, Misset B, Dupuis C, Veber B, Debarre M, Collange O, Pottecher J, Hecketsweiler S, Fromentin M, Tesnière A, Koch C, Sander M, Elke G, Wrigge H, Simon P, Chalkiadaki A, Tzanidakis C, Pneumatikos I, Sertaridou E, Mastora Z, Pantazopoulos I, Papanikolaou M, Papavasilopoulou T, Floros J, Kolonia V, Diakaki C, Rallis M, Paridou A, Kalogeromitros A, Romanou V, Nikolaou C, Kounougeri K, Tsigou E, Psallida V, Karampela N, Mandragos K, Kontoudaki E, Pentheroudaki A, Farazi-Chongouki C, Karakosta A, Chouris I, Radu V, Malliotakis P, Kokkini S, Charalambous E, Kyritsi A, Koulouras V, Papathanakos G, Naglry E, Lampiri C, Tsimpoukas F, Sarakatsanos I, Georgakopoulos P, Ravani I, Prekates A, Sakellaridis K, Christopoulos C, Vrettou Ef, Stokkos K, Pentari A, Marmanidou K, Kydona C, Tsoumaropoulos G, Bitzani M, Kontou P, Voudouris A, Elli N, Flioni, Antypa E, Chasou E, Anisoglou S, Papageorgiou E, Paraforou T, Tsioka A, Karathanou A, Vakalos A, Shah B, Thakkar C, Jain N, Gurjar M, Baronia A, Sathe P, Kulkarni S, Paul C, Paul J, Masjedi M, Nikandish R, Zand F, Sabetian G, Mahmoodpoor A, Hashemian SM, Bala M, Flocco R, Torrente S, Pota V, Spadaro S, Volta C, Serafini G, Boraso S, Tiberio I, Cortegiani A, Misseri G, Barbagallo M, Nicolotti D, Forfori F, Corradi F, De Pascale G, Pelagalli L, Brazzi L, Vittone FG, Russo A, Simion D, Cotoia A, Cinnella G, Toppin P, Johnson-Jackson R, Hayashi Y, Yamamoto R, Yasuda H, Kishihara Y, Shiotsuka J, Sanchez-Hurtado LA, Tejeda-Huezo B, Gorordo L, Ñamendys-Silva SA, Garcia-Guillen FJ, Martinez M, Romero-Meja E, Colorado-Dominguez E, van den Oever H, Kalff KM, Vermeijden W, Cornet AD, Beck O, Cimic N, Dormans T, Bormans L, Bakker J, Van Duijn D, Bosman G, Vos P, Kesecioglu J, Haas L, Henein A, Miranda AM, Malca GEG, Arroyo-Sanchez A, Misiewska-Kaczur A, Akinyi F, Czuczwar M, Luczak K, Sulkowski W, Tamowicz B, Swit B, Baranowski B, Smuszkiewicz P, Trojanowska I, Rzymski S, Sawinski M, Trosiak M, Mikaszewska-Sokolewicz M, Alves R, Leal D, Krystopchuk A, Mendonca PMH, Pereira RA, de Carvalho MRLM, Candeias C, Molinos E, Ferreira A, Castro G, Pereira J-M, Santos L, Ferreira A, Pascoalinho D, Ribeiro R, Domingos G, Gomes P, Nora D, Costa RP, Santos A, Alsheikhly AS, Popescu M, Grigoras I, Patrascanu E, Zabolotskikh I, Musaeva T, Gaigolnik D, Kulabukhov V, Belskiy V, NadezhdaZubareva, Tribulev M, Abdelsalam A, Aldarsani A, Al-Khalid M, Almekhlafi G, Mandourah Y, Doklestic K, Velickovic J, Velickovic D, Jankovic R, Skoric-Jokic S, Radovanovic D, Richards G, Alli A, Nielfa MdCC, Iniesta RS, Martínez AB-C, Bernedo CG, Gil SAP, Nuvials X, Garcia JG, Peña JMG, Jimenez R, Herrera L, Barrachina LG, Monzon IC, Redondo FJ, Villazala R, Zapata DFM, Lopez IMV, Moreno-Gonzalez G, Lopez-Delgado JC, Marin JS, Sanchez-Zamora P, Vidal MV, Gonzalez JF, Salinas I, Hermosa C, Martinez-Sagasti F, Domingo-Marín S, Victorino JA, Garcia-Alvarez R, Calleja PL-A, de la Torre-Prados M-V, Vidal-Cortes P, del Río-Carbajo L, Izura J, Minguez V, Alvarez JT, Prous AP, Paz D, Roche-Campo F, Aguilar G, Belda J, Rico-Feijoo J, Aldecoa C, Zalba-Etayo B, Lang M, Dullenkopf A, Trongtrakul K, Chtsomkasem A, Akbas T, Unal MN, Ozcelik M, Gumus A, Ramazanoglu A, Memis D, Mehmet I, Urkmez S, Ozgultekin A, Demirkiran O, Aslan NA, Kizilaslan D, Kahveci F, Ünlü N, Ozkan Z, Kaye C, Jansen J, O'Neill O, Nutt C, Jha R, Hooker N, Grecu I, Petridou C, Shyamsundar M, McNamee L, Trinder J, Hagan S, Kelly C, Silversides J, Groba CB, Boyd O, Bhowmick K, Humphreys S, Summers C, Polgarova P, Margarson M, Dickens J, Pearson S, Chinery E, Hemmings N, O'Kane S, Austin P, Cole S, Plowright C, Box R, Wright C, Young L, Montague L, Parker R, Morton B, Ostermann M, Bilinska J, Rose BO, Reece-Anthony R, Ryan C, Hamilton M, Hopkins P, Wendon J, Brescia G, Ijaz N, Wood J, George M, Toth-Tarsoly P, Yates B, Armstrong M, Scott C, Boyd C, Szakmany T, Rees D, Pulak P, Coggon M, Saha B, Kent L, Gibson B, Camsooksai J, Reschreiter H, Morgan P, Sangaralingham S, Lowe A, Vondras P, Jamadarkhana S, Cruz C, Bhandary R, Hersey P, Furneval J, Innes R, Doble P, Attwood B, Parsons P, Page V, Zhao X, Dalton J, Hegazy M, Awad Y, Naylor D, Naylor A, Lee S, Brevard S, Davis N, European Soc Intensive Care M. Epidemiology of intra-abdominal infection and sepsis in critically ill patients: "AbSeS", a multinational observational cohort study and ESICM Trials Group Project. INTENSIVE CARE MEDICINE 2019; 45:1703-17.

[607] Chen SL, Ding Y, Apisarnthanarak A, Kalimuddin S, Archuleta S, Omar SFS, De PP, Koh TH, Chew KL, Atiya N, Suwantarat N, Velayuthan RD, Wong JGX, Lye DCB. The higher prevalence of extended spectrum beta-lactamases

among Escherichia coli ST131 in Southeast Asia is driven by expansion of a single, locally prevalent subclone. Sci Rep 2019; 9:13245.
[608] Hendriksen RS, Munk P, Njage P, van Bunnik B, McNally L, Lukjancenko O, Röder T, Nieuwenhuijse D, Pedersen SK, Kjeldgaard J, Kaas RS, Clausen PTLC, Vogt JK, Leekitcharoenphon P, van de Schans MGM, Zuidema T, de Roda Husman AM, Rasmussen S, Petersen B, Bego A, Rees C, Cassar S, Coventry K, Collignon P, Allerberger F, Rahube TO, Oliveira G, Ivanov I, Vuthy Y, Sopheak T, Yost CK, Ke C, Zheng H, Li B, Jiao X, Donado-Godoy P, Coulibaly KJ, Jergović M, Hrenovic J, Karpíšková R, Villacis JE, Legesse M, Eguale T, Heikinheimo A, Malania L, Nitsche A, Brinkmann A, Saba CKS, Kocsis B, Solymosi N, Thorsteinsdottir TR, Hatha AM, Alebouyeh M, Morris D, Cormican M, O'Connor L, Moran-Gilad J, Alba P, Battisti A, Shakenova Z, Kiiyukia C, Ng'eno E, Raka L, Avsejenko J, Bērziņš A, Bartkevics V, Penny C, Rajandas H, Parimannan S, Haber MV, Pal P, Jeunen G-J, Gemmell N, Fashae K, Holmstad R, Hasan R, Shakoor S, Rojas MLZ, Wasyl D, Bosevska G, Kochubovski M, Radu C, Gassama A, Radosavljevic V, Wuertz S, Zuniga-Montanez R, Tay MYF, Gavačová D, Pastuchova K, Truska P, Trkov M, Esterhuyse K, Keddy K, Cerdà-Cuéllar M, Pathirage S, Norrgren L, Örn S, Larsson DGJ, van der Heijden T, Kumburu HH, Sanneh B, Bidjada P, Njanpop-Lafourcade B-M, Nikiema-Pessinaba SC, Levent B, Meschke JS, Beck NK, Van CD, Phuc ND, Tran DMN, Kwenda G, Tabo D-a, Wester AL, Cuadros-Orellana S, Amid C, Cochrane G, Sicheritz-Ponten T, Schmitt H, Alvarez JRM, Aidara-Kane A, Pamp SJ, Lund O, Hald T, Woolhouse M, Koopmans MP, Vigre H, Petersen TN, Aarestrup FM, The Global Sewage Surveillance project c. Global monitoring of antimicrobial resistance based on metagenomics analyses of urban sewage. Nature Communications 2019; 10:1124.
[609] Hsia Y, Lee BR, Versporten A, Yang Y, Bielicki JA, Jackson C, Newland J, Goossens H, Magrini N, Sharland M. Use of the WHO Access, Watch, and Reserve classification to define patterns of hospital antibiotic use (AWaRe): an analysis of paediatric survey data from 56 countries. Lancet Glob Health 2019; 7:e861-e71.
[610] Hsia Y, Sharland M, Jackson C, Wong ICK, Magrini N, Bielicki JA. Consumption of oral antibiotic formulations for young children according to the WHO Access, Watch, Reserve (AWaRe) antibiotic groups: an analysis of sales data from 70 middle-income and high-income countries. Lancet Infect Dis 2019; 19:67-75.
[611] Hu YJ, Ogyu A, Cowling BJ, Fukuda K, Pang HH. Available evidence of antibiotic resistance from extended-spectrum β-lactamase-producing Enterobacteriaceae in paediatric patients in 20 countries: a systematic review and meta-analysis. Bulletin of the World Health Organization 2019; 97:486-501B.
[612] Luo Y, Cheong E, Bian Q, Collins DA, Ye J, Shin JH, Yam W-C, Takata T, Song X, Wang X, Kamboj M, Gottlieb T, Jiang J, Riley TV, Tang Y-W, Jin D. Different molecular characteristics and antimicrobial resistance profiles of Clostridium difficile in the Asia-Pacific region. Emerg Microbes Infect 2019; 8:1553-62.
[613] Ma L, Li B, Zhang T. New insights into antibiotic resistome in drinking water and management perspectives: A metagenomic based study of small-sized microbes. Water Res 2019; 152:191-201.
[614] Papadopoulos A, Ribera A, Mavrogenis AF, Rodriguez-Pardo D, Bonnet E, Salles MJ, Del Toro MD, Nguyen S, Blanco-García A, Skaliczki G, Soriano A, Benito N, Petersdorf S, Pasticci MB, Tattevin P, Tufan ZK, Chan M, O'Connell N, Pantazis N, Kyprianou A, Pigrau C, Megaloikonomos PD, Senneville E, Ariza J, Papagelopoulos PJ, Giannitsioti E. Multidrug-resistant and extensively drug-resistant Gram-negative prosthetic joint infections: Role of surgery and impact of colistin administration. Int J Antimicrob Agents 2019; 53:294-301.
[615] Cervantes S, Prudhomme J, Carter D, Gopi KG, Li Q, Chang Y-T, Le Roch KG. High-content live cell imaging with RNA probes: advancements in high-throughput antimalarial drug discovery. BMC CELL BIOLOGY 2009; 10.
[616] Tam VH, Ledesma KR, Schilling AN, Lim T-P, Yuan Z, Ghose R, Lewis RE. In vivo dynamics of carbapenem-resistant Pseudomonas aeruginosa selection after suboptimal dosing. DIAGNOSTIC MICROBIOLOGY AND INFECTIOUS DISEASE 2009; 64:427-33.
[617] Valvatne H, Syre H, Kross M, Stavrum R, Ti T, Phyu S, Grewal HM. Isoniazid and rifampicin resistance-associated mutations in Mycobacterium tuberculosis isolates from Yangon, Myanmar: implications for rapid molecular testing. JOURNAL OF ANTIMICROBIAL CHEMOTHERAPY 2009; 64:694-701.
[618] Dhorda M, Nyehangane D, Rénia L, Piola P, Guirin PJ, Snounou G. Transmission of Plasmodium vivax in South-Western Uganda: Report of Three Cases in Pregnant Women. PLOS ONE 2011; 6.
[619] Leitsch D, Burgess AG, Dunn LA, Krauer KG, Tan K, Duchene M, Upcroft P, Eckmann L, Upcroft JA. Pyruvate:ferredoxin oxidoreductase and thioredoxin reductase are involved in 5-nitroimidazole activation while flavin metabolism is linked to 5-nitroimidazole resistance in Giardia lamblia. J Antimicrob Chemother 2011; 66:1756-65.
[620] Massi MN, Wahyuni S, Halik H, (NITD) A, Yusuf I, Leong F, Dick T, Phyu S, Massi MN. Drug resistance among tuberculosis patients attending diagnostic and treatment centres in Makassar, Indonesia. International Journal of Tuberculosis and Lung Disease 2011; 15:489-95.
[621] Siswantoro H, Russell B, Ratcliff A, Prasetyorini B, Chalfein F, Marfurt J, Kenangalem E, Wuwung M, Piera KA, Ebsworth E, Anstey NM, Tjitra E, Price RN. In Vivo and In Vitro Efficacy of Chloroquine against Plasmodium malariae and P. ovale in Papua, Indonesia. ANTIMICROBIAL AGENTS AND CHEMOTHERAPY 2011; 55:197-202.

[622] Apisarnthanarak A, Hsu L-Y, Warren DK. Termination of an Extreme-Drug Resistant-Acinetobacter baumannii Outbreak in a Hospital After Flooding: Lessons Learned. Clin Infect Dis 2012; 55:1589-90.

[623] Brunner R, Aissaoui H, Boss C, Bozdech Z, Brun R, Corminboeuf O, Delahaye S, Fischli C, Heidmann B, Kaiser M, Kamber J, Meyer S, Papastogiannidis P, Siegrist R, Voss TS, Welford R, Wittlin S, Binkert C. Identification of a New Chemical Class of Antimalarials. JOURNAL OF INFECTIOUS DISEASES 2012; 206:735-43.

[624] Cervantes S, Stout PE, Prudhomme J, Engel S, Bruton M, Cervantes M, Carter D, Chang Y-T, Hay ME, Aalbersberg W, Kubanek J, Le Roch KG. High content live cell imaging for the discovery of new antimalarial marine natural products. BMC INFECTIOUS DISEASES 2012; 12.

[641] Yepuri NR, Barraud N, Mohammadi NS, Kardak BG, Kjelleberg S, Rice SA, Kelso MJ. Synthesis of cephalosporin-3′-diazeniumdiolates: Biofilm dispersing NO-donor prodrugs activated by β-lactamase. Chemical Communications 2013; 49:4791-3.
[642] Zhang Y, Parker DB, Snow DD, Zhou Z, Li X. Intracellular and extracellular antimicrobial resistance genes in the sludge of livestock waste management structures. Environmental Science and Technology 2013; 47:10206-13.
[643] Zhang Y, Zhang C, Parker DB, Snow DD, Zhou Z, Li X. Occurrence of antimicrobials and antimicrobial resistance genes in beef cattle storage ponds and swine treatment lagoons. Science of the Total Environment 2013; 463-464:631-8.
[644] Apisarnthanarak A, Hsu L-Y, Lim T-P, Mundy LM. Increase in chlorhexidine minimal inhibitory concentration of Acinetobacter baumannii clinical isolates after implementation of advanced source control. Infection Control and Hospital Epidemiology 2014; 35:98-9.
[645] Butler J, Hooper KA, Petrie S, Lee RTC, Maurer-Stroh S, Reh L, Guarnaccia T, Baas C, Xue L, Vitesnik S, Leang S-K, McVernon J, Kelso A, Barr IG, McCaw JM, Bloom JD, Hurt AC. Estimating the fitness advantage conferred by permissive neuraminidase mutations in recent oseltamivir-resistant A(H1N1)pdm09 influenza viruses. PLoS Pathog 2014; 10:e1004065.
[646] Duan S, Govorkova EA, Bahl J, Zaraket H, Baranovich T, Seiler P, Prevost K, Webster RG, Webby RJ. Epistatic interactions between neuraminidase mutations facilitated the emergence of the oseltamivir-resistant H1N1 influenza viruses. Nat Commun 2014; 5:5029.
[647] Karunakaran R, Tay ST, Rahim FF, Lim BB, Puthucheary SD. Molecular Analysis of Ciprofloxacin Resistance among Non-Typhoidal Salmonella with Reduced Susceptibility to Ciprofloxacin Isolated from Patients at a Tertiary Care Hospital in Kuala Lumpur, Malaysia. JAPANESE JOURNAL OF INFECTIOUS DISEASES 2014; 67:157-62.
[648] Kelesidis T, Chow ALP. Proximity to animal or crop operations may be associated with de novo daptomycin-non-susceptible Enterococcus infection. EPIDEMIOLOGY AND INFECTION 2014; 142:221-4.
[649] Lai C-c, Lee K, Xiao Y, Ahmad N, Veeraraghavan B, Thamlikitkul V, Tambyah PA, Nelwan R, Shibl AM, Wu J-J, Seto W-H, Hsueh P-R. High burden of antimicrobial drug resistance in Asia. J Glob Antimicrob Resist 2014; 2:141-7.
[650] Landelle C, Marimuthu K, Harbarth S. Infection control measures to decrease the burden of antimicrobial resistance in the critical care setting. Curr Opin Crit Care 2014; 20:499-506.
[651] Stryjewski ME, Lentnek A, O'Riordan W, Pullman J, Tambyah PA, Miró JM, Fowler Jr VG, Barriere SL, Kitt MM, Corey GR. A randomized Phase 2 trial of telavancin versus standard therapy in patients with uncomplicated Staphylococcus aureus bacteremia: the ASSURE study. BMC INFECTIOUS DISEASES 2014; 14.
[652] Veiga MI, Osório NS, Ferreira PE, Franzén O, Dahlstrom S, Lum JK, Nosten FH, Gil JP. Complex polymorphisms in the Plasmodium falciparum multidrug resistance protein 2 gene and its contribution to antimalarial response. Antimicrob Agents Chemother 2014; 58:7390-7.
[653] Aamodt H, Mohn SC, Maselle S, Manji KP, Willems R, Jureen R, Langeland N, Blomberg B. Genetic relatedness and risk factor analysis of ampicillin-resistant and high-level gentamicin-resistant enterococci causing bloodstream infections in Tanzanian children. BMC Infect Dis 2015; 15:107.
[654] Baranovich T, Bahl J, Marathe BM, Culhane M, Stigger-Rosser E, Darnell D, Kaplan BS, Lowe JF, Webby RJ, Govorkova EA. Influenza A viruses of swine circulating in the United States during 2009-2014 are susceptible to neuraminidase inhibitors but show lineage-dependent resistance to adamantanes. Antiviral Res 2015; 117:10-9.
[655] Farrukee R, Leang S-K, Butler J, Lee RTC, Maurer-Stroh S, Tilmanis D, Sullivan S, Mosse J, Barr IG, Hurt AC. Influenza viruses with B/Yamagata- and B/Victoria-like neuraminidases are differentially affected by mutations that alter antiviral susceptibility. J Antimicrob Chemother 2015; 70:2004-12.
[656] Malmquist NA, Sundriyal S, Caron J, Chen P, Witkowski B, Menard D, Suwanarusk R, Rénia L, Nosten FH, Jiménez-Díaz MB, Angulo-Barturen I, Martínez MS, Ferrer S, Sanz LM, Gamo F-J, Wittlin S, Duffy S, Avery VM, Ruecker A, Delves MJ, Sinden RE, Fuchter MJ, Scherf A. Histone methyltransferase inhibitors are orally bioavailable, fast-acting molecules with activity against different species causing malaria in humans. Antimicrob Agents Chemother 2015; 59:950-9.
[657] Matsugana S, Masaoka T, Sawasaki T, Morishita R, Iwantani Y, Tatsumi M, Endo Y, Yamamoto N, Sugiura W, Ryo A. A cell-free enzymatic activity assay for the evaluation of HIV-1 drug resistance to protease inhibitors. Front Microbiol 2015; 6:1220.
[658] Nguyen D, Nguyen T-K, Rice SA, Boyer C. CO-Releasing Polymers Exert Antimicrobial Activity. Biomacromolecules 2015; 16:2776-86.
[659] Regmi SM, Chaiprasert A, Coker OO, Disratthakit A, Prammananan T, Suriyaphol P, Teo Y-Y, Ong RT-H. Draft Genome Sequence of an Extensively Drug-Resistant Mycobacterium tuberculosis Manu-Ancestor Spoligo-International Type 523 Isolate from Thailand. Genome Announc 2015; 3:e01589-14.

[660] Rhee S-Y, Jordan MR, Raizes E, Chua A, Parkin N, Kantor R, Van Zyl GU, Mukui I, Hosseinipour MC, Frenkel LM, Ndembi N, Hamers RL, De Wit TFR, Wallis CL, Gupta RK, Fokam J, Zeh C, Schapiro JM, Carmona S, Katzenstein D, Tang M, Aghokeng AF, De Oliveira T, Wensing AM, Gallant JE, Wainberg MA, Richman DD, Fitzgibbon JE, Schito M, Bertagnolio S, Yang C, Shafer RW. HIV-1 Drug Resistance Mutations: Potential Applications for Point-of-Care Genotypic Resistance Testing. PLoS One 2015; 10:e0145772.
[661] Soetaert K, Rens C, Wang X-M, De Bruyn J, Lanéelle M-A, Laval F, Lemassu A, Daffé M, Bifani P, Fontaine V, Lefèvre P. Increased Vancomycin Susceptibility in Mycobacteria: a New Approach To Identify Synergistic Activity against Multidrug-Resistant Mycobacteria. Antimicrob Agents Chemother 2015; 59:5057-60.
[662] Zowawi HM, Harris PNA, Roberts MJ, Tambyah PA, Schembri MA, Pezzani MD, Williamson DA, Paterson DL. The emerging threat of multidrug-resistant Gram-negative bacteria in urology. Nat Rev Urol 2015; 12:570-84.
[663] Arribas JR, Girard P-M, Paton NI, Winston A, Marcelin A-G, Elbirt D, Hill A, Hadacek MB. Efficacy of protease inhibitor monotherapy vs. triple therapy: meta-analysis of data from 2303 patients in 13 randomized trials. HIV Med 2016; 17:358-67.
[664] Auburn S, Serre D, Pearson RD, Amato R, Sriprawat K, To S, Handayuni I, Suwanarusk R, Russell B, Drury E, Stalker J, Miotto O, Kwiatkowski DP, Nosten FH, Price RN. Genomic Analysis Reveals a Common Breakpoint in Amplifications of the Plasmodium vivax Multidrug Resistance 1 Locus in Thailand. J Infect Dis 2016; 214:1235-42.
[665] Coker OO, Chaiprasert A, Ngamphiw C, Tongsima S, Regmi SM, Clark TG, Ong RT-H, Teo Y-Y, Prammananan T, Palittapongarnpim P. Genetic signatures of Mycobacterium tuberculosis Nonthaburi genotype revealed by whole genome analysis of isolates from tuberculous meningitis patients in Thailand. PeerJ 2016; 4:e1905.
[666] Cunningham SA, Vasoo S, Patel R. Evaluation of the Check-Points Check MDR CT103 and CT103 XL Microarray Kits by Use of Preparatory Rapid Cell Lysis. J Clin Microbiol 2016; 54:1368-71.
[667] Grigg MJ, William T, Menon J, Barber BE, Wilkes CS, Rajahram GS, Edstein MD, Auburn S, Price RN, Yeo TW, Anstey NM. Efficacy of Artesunate-mefloquine for Chloroquine-resistant Plasmodium vivax Malaria in Malaysia: An Open-label, Randomized, Controlled Trial. Clin Infect Dis 2016; 62:1403-11.
[668] Hanafi A, Lee WC, Loke MF, Teh X, Shaari A, Dinarvand M, Lehours P, Mégraud F, Leow AHR, Vadivelu J, Goh KL. Molecular and Proteomic Analysis of Levofloxacin and Metronidazole Resistant Helicobacter pylori. Front Microbiol 2016; 7:2015.
[669] Harris RC, Khan MS, Martin LJ, Allen V, Moore DAJ, Fielding K, Grandjean L. The effect of surgery on the outcome of treatment for multidrug-resistant tuberculosis: a systematic review and meta-analysis. BMC Infect Dis 2016; 16:262.
[670] Mac Aogáin M, Miajlovic H, Moloney G, Chotirmall SH, Rogers TR, Smith SGJ. Identification of a novel sequence type of Escherichia coli as the causative agent of pyelonephritis and bloodstream infection. JMM Case Rep 2016; 3:e005061.
[671] Nguyen T-K, Selvanayagam R, Ho KKK, Chen R, Kutty SK, Rice SA, Kumar N, Barraud N, Duong HTT, Boyer C. Co-delivery of nitric oxide and antibiotic using polymeric nanoparticles. Chem Sci 2016; 7:1016-27.
[672] Nilsson M, Rybtke MT, Givskov M, Høiby N, Twetman S, Tolker-Nielsen T. The dlt genes play a role in antimicrobial tolerance of Streptococcus mutans biofilms. INTERNATIONAL JOURNAL OF ANTIMICROBIAL AGENTS 2016; 48:298-304.
[673] Phyo AP, Ashley EA, Anderson TJC, Bozdech Z, Carrara VI, Sriprawat K, Nair S, White MM, Dziekan J, Ling C, Proux S, Konghahong K, Jeeyapant A, Woodrow CJ, Imwong M, McGready R, Lwin KM, Day NPJ, White NJ, Nosten FH. Declining Efficacy of Artemisinin Combination Therapy Against P. Falciparum Malaria on the Thai-Myanmar Border (2003-2013): The Role of Parasite Genetic Factors. Clin Infect Dis 2016; 63:784-91.
[674] Richmond GE, Evans LP, Anderson MJ, Wand ME, Bonney LC, Ivens A, Chua KL, Webber MA, Sutton JM, Peterson ML, Piddock LJV. The Acinetobacter baumannii Two-Component System AdeRS Regulates Genes Required for Multidrug Efflux, Biofilm Formation, and Virulence in a Strain-Specific Manner. mBio 2016; 7:e00430-16.
[675] Stenvang M, Dueholm MS, Vad BS, Seviour T, Zeng G, Geifman-Shochat S, Søndergaard MT, Christiansen G, Meyer RL, Kjelleberg S, Nielsen PH, Otzen DE. Epigallocatechin Gallate Remodels Overexpressed Functional Amyloids in Pseudomonas aeruginosa and Increases Biofilm Susceptibility to Antibiotic Treatment. J Biol Chem 2016; 291:26540-53.
[676] Atarashi K, Suda W, Luo C, Kawaguchi T, Motoo I, Narushima S, Kiguchi Y, Yasuma K, Watanabe E, Tanoue T, Thaiss CA, Sato M, Toyooka K, Said HS, Yamagami H, Rice SA, Gevers D, Johnson RC, Segre JA, Chen K, Kolls JK, Elinav E, Morita H, Xavier RJ, Hattori M, Honda K. Ectopic colonization of oral bacteria in the intestine drives T(H)1 cell induction and inflammation. Science 2017; 358:359-65.
[677] Basilico N, Parapini S, Sparatore A, Romeo S, Misiano P, Vivas L, Yardley V, Croft SL, Habluetzel A, Lucantoni L, Rénia L, Russell B, Suwanarusk R, Nosten FH, Dondio G, Bigogno C, Jabes D, Taramelli D. In Vivo and In Vitro Activities

and ADME-Tox Profile of a Quinolizidine-Modified 4-Aminoquinoline: A Potent Anti-P. falciparum and Anti-P. vivax Blood-Stage Antimalarial. Molecules 2017; 22:2102.
[678] Bazaka K, Bazaka O, Levchenko I, Xu S, Ivanova EP, Keidar M, Ostrikov KK. Plasma-potentiated small molecules—possible alternative to antibiotics? Nano Futures 2017; 1.
[679] Belousoff MJ, Eyal Z, Radjainia M, Ahmed T, Bamert RS, Matzov D, Bashan A, Zimmerman E, Mishra S, Cameron D, Elmlund H, Peleg AY, Bhushan S, Lithgow T, Yonath A. Structural Basis for Linezolid Binding Site Rearrangement in the Staphylococcus aureus Ribosome. mBio 2017; 8.
[680] Bhuyan GS, Hossain MA, Sarker SK, Rahat A, Islam MT, Haque TN, Begum N, Qadri SK, Muraduzzaman AKM, Islam NN, Islam MS, Sultana N, Jony MHK, Khanam F, Mowla G, Matin A, Begum F, Shirin T, Ahmed D, Saha N, Qadri F, Mannoor K. Bacterial and viral pathogen spectra of acute respiratory infections in under-5 children in hospital settings in Dhaka city. PLoS One 2017; 12:e0174488.
[681] Chang MJ, Jin B, Chae J-w, Yun H-y, Kim ES, Lee YJ, Cho Y-J, Yoon HI, Lee C-T, Park KU, Song J, Lee J-H, Park JS. Population pharmacokinetics of moxifloxacin, cycloserine, p-aminosalicylic acid and kanamycin for the treatment of multi-drug-resistant tuberculosis. Int J Antimicrob Agents 2017; 49:677-87.
[682] Chen F, Chen G, Liu Y, Jin Y, Cheng Z, Liu Y, Yang L, Jin S, Wu W. Pseudomonas aeruginosa oligoribonuclease contributes to tolerance to ciprofloxacin by regulating pyocin biosynthesis. Antimicrobial Agents and Chemotherapy 2017; 61.
[683] Cunningham SA, Limbago B, Traczewski M, Anderson K, Hackel MA, Hindler J, Sahm DF, Alyanak E, Lawsin A, Gulvik CA, de Man TJB, Mandrekar JN, Schuetz AN, Jenkins S, Humphries R, Palavecino E, Vasoo S, Patel R. Multicenter Performance Assessment of Carba NP Test. J Clin Microbiol 2017; 55:1954-60.
[684] Gowrisankar G, Chelliah R, Ramakrishnan SR, Elumalai V, Dhanamadhavan S, Brindha K, Antony U, Elango L. Chemical, microbial and antibiotic susceptibility analyses of groundwater after a major flood event in Chennai. Sci Data 2017; 4:170135.
[685] Gunawan C, Marquis CP, Amal R, Sotiriou GA, Rice SA, Harry EJ. Widespread and Indiscriminate Nanosilver Use: Genuine Potential for Microbial Resistance. ACS NANO 2017; 11:3438-45.
[686] Howlin RP, Cathie K, Hall-Stoodley L, Cornelius V, Duignan C, Allan RN, Fernandez BO, Barraud N, Bruce KD, Jefferies JMC, Kelso MJ, Kjelleberg S, Rice SA, Rogers GB, Pink S, Smith C, Sukhtankar PS, Salib R, Legg J, Carroll M, Daniels T, Feelisch M, Stoodley P, Clarke SC, Connett G, Faust SN, Webb JS. Low-Dose Nitric Oxide as Targeted Anti-biofilm Adjunctive Therapy to Treat Chronic Pseudomonas aeruginosa Infection in Cystic Fibrosis. Molecular Therapy 2017; 25:2104-16.
[687] Hutchison C, Khan MS, Yoong J, Lin X, Coker RJ. Financial barriers and coping strategies: a qualitative study of accessing multidrug-resistant tuberculosis and tuberculosis care in Yunnan, China. BMC Public Health 2017; 17:221.
[688] Kathirvel S, Tripathy JP, Tun ZM, Patro BK, Singh T, Bhalla A, Devnani M, Wilkinson E. Physicians' compliance with the National Drug Policy on Malaria in a tertiary teaching hospital, India, from 2010 to 2015: a mixed method study. Trans R Soc Trop Med Hyg 2017; 111:62-70.
[689] Lamoth F, Chung SJ, Damonti L, Alexander BD. Changing Epidemiology of Invasive Mold Infections in Patients Receiving Azole Prophylaxis. Clin Infect Dis 2017; 64:1619-21.
[690] Landier J, Kajeechiwa L, Thwin MM, Parker DM, Chaumeau V, Wiladphaingern J, Imwong M, Miotto O, Patumrat K, Duanguppama J, Cerqueira D, Malleret B, Rénia L, Nosten S, von Seidlein L, Ling C, Proux S, Corbel V, Simpson JA, Dondorp AM, White NJ, Nosten FH. Safety and effectiveness of mass drug administration to accelerate elimination of artemisinin-resistant falciparum malaria: A pilot trial in four villages of Eastern Myanmar. Wellcome Open Res 2017; 2:81.
[691] Liao J-H, Tsai C-H, Patel SG, Yang J-T, Tu I-F, Lo Cicero M, Lipka-Lloyd M, Wu W-L, Shen W-J, Ho M-R, Chou C-C, Sharma GR, Okanishi H, Luk LYP, Tsai Y-H, Wu S-H. Acetylome of Acinetobacter baumannii SK17 Reveals a Highly-Conserved Modification of Histone-Like Protein HU. Front Mol Biosci 2017; 4:77.
[692] Thai VC, Lim TK, Le KPU, Lin Q, Nguyen TTH. iTRAQ-based proteome analysis of fluoroquinolone-resistant Staphylococcus aureus. J Glob Antimicrob Resist 2017; 8:82-9.
[693] Ahmed W, Zhang Q, Lobos A, Senkbeil J, Sadowsky MJ, Harwood VJ, Saeidi N, Marinoni O, Ishii S. Precipitation influences pathogenic bacteria and antibiotic resistance gene abundance in storm drain outfalls in coastal sub-tropical waters. Environ Int 2018; 116:308-18.
[694] Antonoplis A, Zang X, Huttner MA, Chong KKL, Lee YB, Co JY, Amieva MR, Kline KA, Wender PA, Cegelski L. A Dual-Function Antibiotic-Transporter Conjugate Exhibits Superior Activity in Sterilizing MRSA Biofilms and Killing Persister Cells. J Am Chem Soc 2018; 140:16140-51.
[695] Beattie RE, Walsh M, Cruz MC, McAliley LR, Dodgen L, Zheng WU-e, Hristova KR. Agricultural contamination impacts antibiotic resistance gene abundances in river bed sediment temporally. FEMS Microbiol Ecol 2018; 94.

[696] Chen C, Gardete S, Jansen RS, Shetty A, Dick T, Rhee KY, Dartois V. Verapamil Targets Membrane Energetics in Mycobacterium tuberculosis. Antimicrob Agents Chemother 2018; 62.
[697] Allix-Béguec C, Arandjelovic I, Bi L, Beckert P, Bonnet M, Bradley P, Cabibbe AM, Cancino-Muñoz I, Caulfield MJ, Chaiprasert A, Cirillo DM, Clifton DA, Comas I, Crook DW, De Filippo MR, de Neeling H, Diel R, Drobniewski FA, Faksri K, Farhat MR, Fleming J, Fowler P, Fowler TA, Gao Q, Gardy J, Gascoyne-Binzi D, Gibertoni-Cruz A-L, Gil-Brusola A, Golubchik T, Gonzalo X, Grandjean L, He G, Guthrie JL, Hoosdally S, Hunt M, Iqbal Z, Ismail N, Johnston J, Khanzada FM, Khor CC, Kohl TA, Kong C, Lipworth S, Liu Q, Maphalala G, Martinez E, Mathys V, Merker M, Miotto P, Mistry N, Moore DAJ, Murray M, Niemann S, Ong RT-H, Peto TEA, Posey JE, Prammananan T, Pym A, Rodrigues C, Rodrigues M, Rodwell T, Rossolini GM, Padilla ES, Schito M, Shen X, Shendure J, Sintchenko V, Sloutsky A, Smith EG, Snyder M, Soetaert K, Starks AM, Supply P, Suriyapol P, Tahseen S, Tang P, Teo Y-Y, Thuong TNT, Thwaites G, Tortoli E, Omar SV, van Soolingen D, Walker AS, Walker TM, Wilcox M, Wilson DJ, Wyllie D, Yang Y, Zhang H, Zhao Y, Zhu B, The CRyPTIC Consortium and the 100 GP. Prediction of Susceptibility to First-Line Tuberculosis Drugs by DNA Sequencing. NEW ENGLAND JOURNAL OF MEDICINE 2018; 379:1403-15.
[698] Dunn DT, Stöhr W, Arenas-Pinto A, Tostevin A, Mbisa JL, Paton NI. Next generation sequencing of HIV-1 protease in the PIVOT trial of protease inhibitor monotherapy. J Clin Virol 2018; 101:63-5.
[699] Fang T, Wang HC-e, Cui Q, Rogers M, Dong P. Diversity of potential antibiotic-resistant bacterial pathogens and the effect of suspended particles on the spread of antibiotic resistance in urban recreational water. Water Res 2018; 145:541-51.
[700] Germond A, Ichimura T, Horinouchi T, Fujita H, Furusawa C, Watanabe TM. Raman spectral signature reflects transcriptomic features of antibiotic resistance in Escherichia coli. Commun Biol 2018; 1:85.
[701] Grigg MJ, William T, Piera KA, Rajahram GS, Jelip J, Aziz A, Menon J, Marfurt J, Price RN, Auburn S, Barber BE, Yeo TW, Anstey NM. Plasmodium falciparum artemisinin resistance monitoring in Sabah, Malaysia: in vivo therapeutic efficacy and kelch13 molecular marker surveillance. Malar J 2018; 17:463.
[702] Hoppe A, Giuliano M, Lugemwa A, Thompson JA, Floridia M, Walker AS, Senoga I, Abwola MC, Pirillo MF, Kityo CM, Arenas-Pinto A, Paton NI. HIV-1 viral load and resistance in genital secretions in patients taking protease-inhibitor-based second-line therapy in Africa. Antivir Ther 2018; 23:191-5.
[703] Jiang Y, Xu C, Wu X, Chen Y, Han W, Gin KY-H, He Y. Occurrence, seasonal variation and risk assessment of antibiotics in Qingcaosha reservoir. Water (Switzerland) 2018; 10.
[704] Kano R, Hsiao Y-H, Han HS, Chen C, Hasegawa A, Kamata H. Resistance Mechanism in a Terbinafine-Resistant Strain of Microsporum canis. Mycopathologia 2018; 183:623-7.
[705] Malkawi R, Iyer A, Parmar A, Lloyd DG, Goh ETL, Taylor EJ, Sarmad S, Madder A, Lakshminarayanan R, Singh I. Cysteines and Disulfide-Bridged Macrocyclic Mimics of Teixobactin Analogues and Their Antibacterial Activity Evaluation against Methicillin-Resistant Staphylococcus Aureus (MRSA). Pharmaceutics 2018; 10:183.
[706] Mather AE, Phuong TLT, Gao Y, Clare S, Mukhopadhyay S, Goulding DA, Hoang NTD, Tuyen HT, Lan NPH, Thompson CN, Trang NHT, Carrique-Mas J, Tue NT, Campbell JI, Rabaa MA, Thanh DP, Harcourt K, Hoa NT, Trung NV, Schultsz C, Perron GG, Coia JE, Brown DJ, Okoro C, Parkhill J, Thomson NR, Chau NVV, Thwaites GE, Maskell DJ, Dougan G, Kenney LJ, Baker S. New Variant of Multidrug-Resistant Salmonella enterica Serovar Typhimurium Associated with Invasive Disease in Immunocompromised Patients in Vietnam. mBio 2018; 9:e01056-18.
[707] Merchant S, Proudfoot EM, Quadri HN, McElroy HJ, Wright WR, Gupta A, Sarpong EM. Risk factors for Pseudomonas aeruginosa infections in Asia-Pacific and consequences of inappropriate initial antimicrobial therapy: A systematic literature review and meta-analysis. J Glob Antimicrob Resist 2018; 14:33-44.
[708] Nordström R, Nyström L, Andrén OCJ, Malkoch M, Umerska A, Davoudi M, Schmidtchen A, Malmsten M. Membrane interactions of microgels as carriers of antimicrobial peptides. J Colloid Interface Sci 2018; 513:141-50.
[709] Oonsivilai M, Mo Y, Luangasanatip N, Lubell Y, Miliya T, Tan P, Loeuk L, Turner P, Cooper BS. Using machine learning to guide targeted and locally-tailored empiric antibiotic prescribing in a children's hospital in Cambodia. Wellcome Open Res 2018; 3:131.
[710] Ravensdale JT, Xian DTW, Wei CM, Lv Q, Wen X, Guo J, Coorey R, LeSouëf P, Lu F, Zhang G, Dykes GA. PCR screening of antimicrobial resistance genes in faecal samples from Australian and Chinese children. J Glob Antimicrob Resist 2018; 14:178-81.
[711] Stockdale AJ, Saunders MJ, Boyd MA, Bonnett LJ, Johnston V, Wandeler G, Schoffelen AF, Ciaffi L, Stafford K, Collier AC, Paton NI, Geretti AM. Effectiveness of Protease Inhibitor/Nucleos(t)ide Reverse Transcriptase Inhibitor-Based Second-line Antiretroviral Therapy for the Treatment of Human Immunodeficiency Virus Type 1 Infection in Sub-Saharan Africa: A Systematic Review and Meta-analysis. Clin Infect Dis 2018; 66:1846-57.
[712] Subedi D, Vijay AK, Kohli GS, Rice SA, Willcox MDP. Association between possession of ExoU and antibiotic resistance in Pseudomonas aeruginosa. PLoS One 2018; 13:e0204936.

[713] Subedi D, Vijay AK, Kohli GS, Rice SA, Willcox MDP. Comparative genomics of clinical strains of Pseudomonas aeruginosa strains isolated from different geographic sites. Sci Rep 2018; 8:15668.
[714] Subedi D, Vijay AK, Kohli GS, Rice SA, Willcox MDP. Nucleotide sequence analysis of NPS-1 β-lactamase and a novel integron (In1427)-carrying transposon in an MDR Pseudomonas aeruginosa keratitis strain. J Antimicrob Chemother 2018; 73:1724-6.
[715] Tzou PL, Ariyaratne P, Varghese V, Lee C, Rakhmanaliev E, Villy C, Yee M, Tan K, Michel G, Pinsky BA, Shafer RW. Comparison of an In Vitro Diagnostic Next-Generation Sequencing Assay with Sanger Sequencing for HIV-1 Genotypic Resistance Testing. J Clin Microbiol 2018; 56.
[716] Vente A, Bentley C, Lückermann M, Tambyah PA, Dalhoff A. Early Clinical Assessment of the Antimicrobial Activity of Finafloxacin Compared to Ciprofloxacin in Subsets of Microbiologically Characterized Isolates. Antimicrob Agents Chemother 2018; 62.
[717] Zhou C, Song H, Loh CJL, She J, Deng L, Liu B. Grafting antibiofilm polymer hydrogel film onto catheter by SARA SI-ATRP. J Biomater Sci Polym Ed 2018; 29:2106-23.
[718] Brunton LA, Desbois AP, Garza M, Wieland B, Mohan CV, Häsler B, Tam CC, Le PNT, Phuong NT, Van PT, Nguyen-Viet H, Eltholth MM, Pham DK, Duc PP, Linh NT, Rich KM, Mateus ALP, Hoque MA, Ahad A, Khan MNA, Adams A, Guitian J. Identifying hotspots for antibiotic resistance emergence and selection, and elucidating pathways to human exposure: Application of a systems-thinking approach to aquaculture systems. Sci Total Environ 2019; 687:1344-56.
[719] Çapcı A, Lorion MM, Wang HD, Simon N, Leidenberger M, Silva MCB, Moreira DRM, Zhu Y, Meng Y, Chen JY, Lee YM, Friedrich O, Kappes B, Wang J, Ackermann L, Tsogoeva SB. Artemisinin-(Iso)quinoline Hybrids by C-H Activation and Click Chemistry: Combating Multidrug-Resistant Malaria. Angew Chem Int Ed Engl 2019; 58:13066-79.
[720] Chen Y, Li P, Huang Y, Yu K, Chen H-J, Cui K, Huang Q, Zhang J, Gin KY-H, He Y. Environmental media exert a bottleneck in driving the dynamics of antibiotic resistance genes in modern aquatic environment. Water Res 2019; 162:127-38.
[721] Chen Y, Su J-Q, Zhang J, Li P, Chen H, Zhang B, Gin KY-H, He Y. High-throughput profiling of antibiotic resistance gene dynamic in a drinking water river-reservoir system. Water Res 2019; 149:179-89.
[722] Faksri K, Kaewprasert O, Ong RT-H, Suriyaphol P, Prammananan T, Teo Y-Y, Srilohasin P, Chaiprasert A. Comparisons of whole-genome sequencing and phenotypic drug susceptibility testing for Mycobacterium tuberculosis causing MDR-TB and XDR-TB in Thailand. Int J Antimicrob Agents 2019; 54:109-16.
[723] González A, Casado J, Chueca E, Salillas S, Velázquez-Campoy A, Angarica VE, Bénejat L, Guignard J, Giese A, Sancho J, Lehours P, Lanas Á. Repurposing Dihydropyridines for Treatment of Helicobacter pylori Infection. Pharmaceutics 2019; 11.
[724] González A, Salillas S, Velázquez-Campoy A, Angarica VE, Fillat MF, Sancho J, Lanas Á. Identifying potential novel drugs against Helicobacter pylori by targeting the essential response regulator HsrA. Sci Rep 2019; 9:11294.
[725] Jabbar A, Phelan JE, de Sessions PF, Khan TA, Rahman H, Khan SN, Cantillon DM, Wildner LM, Ali S, Campino S, Waddell SJ, Clark TG. Whole genome sequencing of drug resistant Mycobacterium tuberculosis isolates from a high burden tuberculosis region of North West Pakistan. Sci Rep 2019; 9:14996.
[726] Juhas M, Widlake E, Teo JWP, Huseby DL, Tyrrell JM, Polikanov YS, Ercan O, Petersson A, Cao S, Aboklaish AF, Rominski A, Crich D, Böttger EC, Walsh TR, Hughes D, Hobbie SN. In vitro activity of apramycin against multidrug-, carbapenem- and aminoglycoside-resistant Enterobacteriaceae and Acinetobacter baumannii. J Antimicrob Chemother 2019; 74:944-52.
[727] Li H, Andersen PS, Stegger M, Sieber RN, Ingmer H, Staubrand N, Dalsgaard A, Leisner JJ. Antimicrobial Resistance and Virulence Gene Profiles of Methicillin-Resistant and -Susceptible Staphylococcus aureus From Food Products in Denmark. Front Microbiol 2019; 10:2681.
[728] Li H, Stegger M, Dalsgaard A, Leisner JJ. Bacterial content and characterization of antibiotic resistant Staphylococcus aureus in Danish sushi products and association with food inspector rankings. Int J Food Microbiol 2019; 305:108244.
[729] Limmathurotsakul D, Sandoe JAT, Barrett DC, Corley M, Hsu L-Y, Mendelson M, Collignon P, Laxminarayan R, Peacock SJ, Howard P. 'Antibiotic footprint' as a communication tool to aid reduction of antibiotic consumption. J Antimicrob Chemother 2019; 74:2122-7.
[730] Long S, Miao L, Li R, Deng F, Qiao Q, Liu X, Yan A, Xu Z. Rapid identification of bacteria by membrane-responsive aggregation of a pyrene derivative. ACS Sensors 2019; 4:281-5.
[731] Nilsson M, Givskov M, Twetman S, Tolker-Nielsen T. Inactivation of the pgmA Gene in Streptococcus mutans Significantly Decreases Biofilm-Associated Antimicrobial Tolerance. Microorganisms 2019; 7.
[732] Nilsson M, Jakobsen TH, Givskov M, Twetman S, Tolker-Nielsen T. Oxidative stress response plays a role in antibiotic tolerance of Streptococcus mutans biofilms. Microbiology (Reading) 2019; 165:334-42.

[733] Pei M, Zhang B, He Y, Su J-Q, Gin KY-H, Lev O, Shen G, Hu S. State of the art of tertiary treatment technologies for controlling antibiotic resistance in wastewater treatment plants. Environ Int 2019; 131:105026.
[734] Penesyan A, Nagy SS, Kjelleberg S, Gillings MR, Paulsen IT. Rapid microevolution of biofilm cells in response to antibiotics. NPJ Biofilms Microbiomes 2019; 5:34.
[735] Phelan JE, Lim DR, Mitarai S, de Sessions PF, Tujan MAA, Reyes LT, Medado IAP, Palparan AG, Naim ANM, Jie S, Segubre-Mercado E, Simoes B, Campino S, Hafalla JC, Murase Y, Morishige Y, Hibberd ML, Kato S, Ama MCG, Clark TG. Mycobacterium tuberculosis whole genome sequencing provides insights into the Manila strain and drug-resistance mutations in the Philippines. Sci Rep 2019; 9:9305.
[736] Ram M R, Teh X, Rajakumar T, Goh KL, Leow AHR, Poh BH, Mariappan V, Shankar EM, Loke MF, Vadivelu J. Polymorphisms in the host CYP2C19 gene and antibiotic-resistance attributes of Helicobacter pylori isolates influence the outcome of triple therapy. J Antimicrob Chemother 2019; 74:11-6.
[737] Safi H, Gopal P, Lingaraju S, Ma S, Levine C, Dartois V, Yee M, Li L, Blanc L, Ho Liang H-P, Husain S, Hoque M, Soteropoulos P, Rustad T, Sherman DR, Dick T, Alland D. Phase variation in Mycobacterium tuberculosis glpK produces transiently heritable drug tolerance. Proc Natl Acad Sci U S A 2019; 116:19665-74.
[738] Sosibo SC, Somboro AM, Amoako DG, Sekyere JO, Bester LA, Ngila JC, Sun DD, Kumalo HM. Impact of Pyridyl Moieties on the Inhibitory Properties of Prominent Acyclic Metal Chelators Against Metallo-β-Lactamase-Producing Enterobacteriaceae: Investigating the Molecular Basis of Acyclic Metal Chelators' Activity. Microb Drug Resist 2019; 25:439-49.
[739] Subedi D, Kohli GS, Vijay AK, Willcox MDP, Rice SA. Accessory genome of the multi-drug resistant ocular isolate of Pseudomonas aeruginosa PA34. PLoS One 2019; 14:e0215038.
[740] Thompson JA, Kityo CM, Dunn DT, Hoppe A, Ndashimye E, Hakim J, Kambugu A, van Oosterhout JJ, Arribas JR, Mugyenyi P, Walker AS, Paton NI. Evolution of Protease Inhibitor Resistance in Human Immunodeficiency Virus Type 1 Infected Patients Failing Protease Inhibitor Monotherapy as Second-line Therapy in Low-income Countries: An Observational Analysis Within the EARNEST Randomized Trial. Clin Infect Dis 2019; 68:1184-92.
[741] Yang DL, Hu YL, Yin ZX, Zeng G, Li D, Zhang YQ, Xu ZH, Guan XM, Weng LX, Wang LH. Cis-2-dodecenoic Acid Mediates Its Synergistic Effect with Triazoles by Interfering with Efflux Pumps in Fluconazole-resistant Candida albicans. Biomed Environ Sci 2019; 32:199-209.
[742] Zhang N, Liu X, Liu R, Zhang T, Li M, Zhang Z, Qu Z, Yuan Z, Yu H. Influence of reclaimed water discharge on the dissemination and relationships of sulfonamide, sulfonamide resistance genes along the Chaobai River, Beijing. FRONTIERS OF ENVIRONMENTAL SCIENCE & ENGINEERING 2019; 13.
